# Enhanced Classification Performance using Deep Learning Based Segmentation for Pulmonary Embolism Detection in CT Angiography

**DOI:** 10.1101/2023.04.21.23288861

**Authors:** Ali Teymur Kahraman, Tomas Fröding, Dimitris Toumpanakis, Christian Jamtheim Gustafsson, Tobias Sjöblom

## Abstract

**Purpose:** To develop a deep learning-based algorithm that automatically and accurately classifies patients as either having pulmonary emboli or not in CT pulmonary angiography (CTPA) examinations.

**Materials and Methods:** For model development, 700 CTPA examinations from 652 patients performed at a single institution were used, of which 149 examinations contained 1497 PE traced by radiologists. The nnU-Net deep learning-based segmentation framework was trained using 5-fold cross-validation. To enhance classification, we applied logical rules based on PE volume and probability thresholds. External model evaluation was performed in 770 and 34 CTPAs from two independent datasets.

**Results:** A total of 1483 CTPA examinations were evaluated. In internal cross-validation and test set, the trained model correctly classified 123 of 128 examinations as positive for PE (sensitivity 96.1%; 95% C.I. 91-98%; *P* < .05) and 521 of 551 as negative (specificity 94.6%; 95% C.I. 92-96%; *P* < .05). In the first external test dataset, the trained model correctly classified 31 of 32 examinations as positive (sensitivity 96.9%; 95% C.I. 84-99%; *P* < .05) and 2 of 2 as negative (specificity 100%; 95% C.I. 34-100%; *P* < .05). In the second external test dataset, the trained model correctly classified 379 of 385 examinations as positive (sensitivity 98.4%; 95% C.I. 97-99%; *P* < .05) and 346 of 385 as negative (specificity 89.9%; 95% C.I. 86-93%; *P* < .05).

**Conclusion:** Our automatic pipeline achieved beyond state-of-the-art diagnostic performance of PE in CTPA using nnU-Net for segmentation and volume- and probability-based post-processing for classification.

**Highlights:**

- An nnU-Net segmentation framework was applied to patient-level classification in CTPA examinations.
- The proposed algorithm can enable prioritization of patients with PE for rapid review in emergency radiology.
- The proposed algorithm showed outstanding performance on both internal and two publicly available external testing datasets (AUC, 98.3%; n=1355).

## 1. Introduction

Pulmonary embolism (PE) is a potentially life-threatening occlusion of pulmonary arteries caused by blood clotting and is associated with significant morbidity and mortality [1]. PE affects >400,000 patients in Europe [2] and between 300,000 and 600,000 patients in the US [3] causing an estimated >100,000 deaths annually [4]. PE is a significant cause of preventable hospital deaths in the world [5], demanding rapid clinical management [6]. The computed tomography pulmonary angiography (CTPA) imaging modality is the current gold standard for PE diagnosis [7]. The CTPA is a CT scan performed after intravenous injection of iodinated contrast medium. As the emboli do not absorb contrast medium they can be recognized as dark filling defects in the pulmonary arteries [8]. Thoroughly examining every CT slice and identification of PE in CTPA is time-consuming for the radiologist and requires considerable training and attentiveness, and the inter-observer variability is high for small, sub-segmental emboli [9]. An automated solution for detection of PE in CTPA has potential to assist the radiologist by reducing reading times and the risk of emboli being overlooked.

Developing a general solution for automatic detection of PE has proven challenging because of anatomical variation, motion and breathing artifacts, inter-patient variability in contrast medium concentration, and concurrent pathologies. Over the past two decades, automated PE detection has been attempted using deterministic models, such as image processing and analysis techniques [10, 11], or probabilistic/statistical models such as machine learning [12–14] and deep convolutional neural networks [15, 16]. Yet, the accuracies of these solutions have been insufficient for clinical use due to low sensitivity [10, 13, 15] and high false positive rate [10, 11, 13, 14], potentially caused by training on small datasets [10, 11, 13–15]. The state-of-the-art is a residual neural network (ResNet) classification architecture on 1465 CTPA examinations with sensitivity of 92.7% and specificity of 95.5% [17]. To mitigate the limited dataset size challenges hampering the training of AI models for PE classification, an alternative approach is to employ a fine-tuned U-Net-like semantic segmentation model. The U-Net model has demonstrated its effectiveness in several medical image segmentation tasks. [18]. The no-new U-Net framework (nnU-Net) successfully addresses challenges of finding the best U-net model and fine-tuning its hyperparameters [19].

Here, we sought to take advantage of the segmentation performance of the nnU-Net framework in an algorithm that automatically classifies routine patient CTPA examinations as having PE or not with higher sensitivity and specificity than the current state-of-the-art performance.

## 2. Materials and Methods

### 2.1. Internal dataset

The single-institution (Nyköping Hospital, Sweden) retrospective dataset consisted of 700 non-ECG-gated CTPA examinations performed between 2014 and 2018 (n=149 positive for PE); 383 CTPA examinations from 353 women (age range 16-97 years; median age 73 years; interquartile range 20 years) and 317 from 299 men (age range 19-100 years; median age 71 years; interquartile range 15 years) [20]. The CTPAs were clinical routine examinations exported in chronological order from a history list in the institution’s Picture Archiving and Communication System (PACS). The only disruption in the order were a few inserted time gaps, which allowed for a larger number of CT scanners to be included as new CT scanners were installed during the time period. The CTPAs were acquired on five different CT scanners (Somatom Definition Flash, Siemens Healthcare, Erlangen, Germany; LightSpeed VCT, General Electric (GE) Healthcare Systems, Waukesha, WI, USA; Brilliance 64, Ingenuity Core and Ingenuity CT, Philips Medical Systems, Eindhoven, the Netherlands). As contrast medium, Omnipaque 350 mg I/ml (GE Healthcare Systems, Waukesha, WI, USA) was used. Collection and analysis of CTPA examinations was approved by the Swedish Ethical Review Authority (EPN Uppsala Dnr 2015/023 and 2015/023/1). The CTPA data was anonymized and exported in Digital Imaging and Communications in Medicine (DICOM) format, using a hardware solution (Dicom2USB). The CTPAs were reviewed and annotated using the open-source software Medical Imaging Interaction Toolkit (MITK) [21] by two radiologists (DT and TF) with 6 and 16 years of experience. Each CTPA was annotated by either DT or TF. All blood clots in 149 CTPA examinations positive for PE were manually segmented in axial view, image by image, resulting in 36,471 segmentations.

### 2.2. External datasets

Two publicly available datasets were used for external evaluation; the Ferdowsi University of Mashhad’s PE dataset (FUMPE) [22] and the RSNA-STR Pulmonary Embolism CT (RSPECT) Dataset [23]. The FUMPE dataset contains 35 CTPAs with voxel-level PE annotation by radiologists. Of the 35 CTPAs, two were negative for PE, 32 were positive and one was excluded for lack of ground truth annotation (Supp. materials). The RSPECT dataset consisted of a training (n=7279) and a test (n=2167) set and image-level annotations were provided for the training set by several subspecialist thoracic radiologists. 385 CTPAs were selected from the RSPECT training dataset out of a total of 398 that had central PE, and 13 examinations were excluded due to errors during the DICOM to NIfTI format conversion process. Of the 4877 CTPAs without PE or other true filling defect, 385 examinations were randomly selected. An overview of our internal and external datasets is shown in Figure 1.

**Figure 1.**
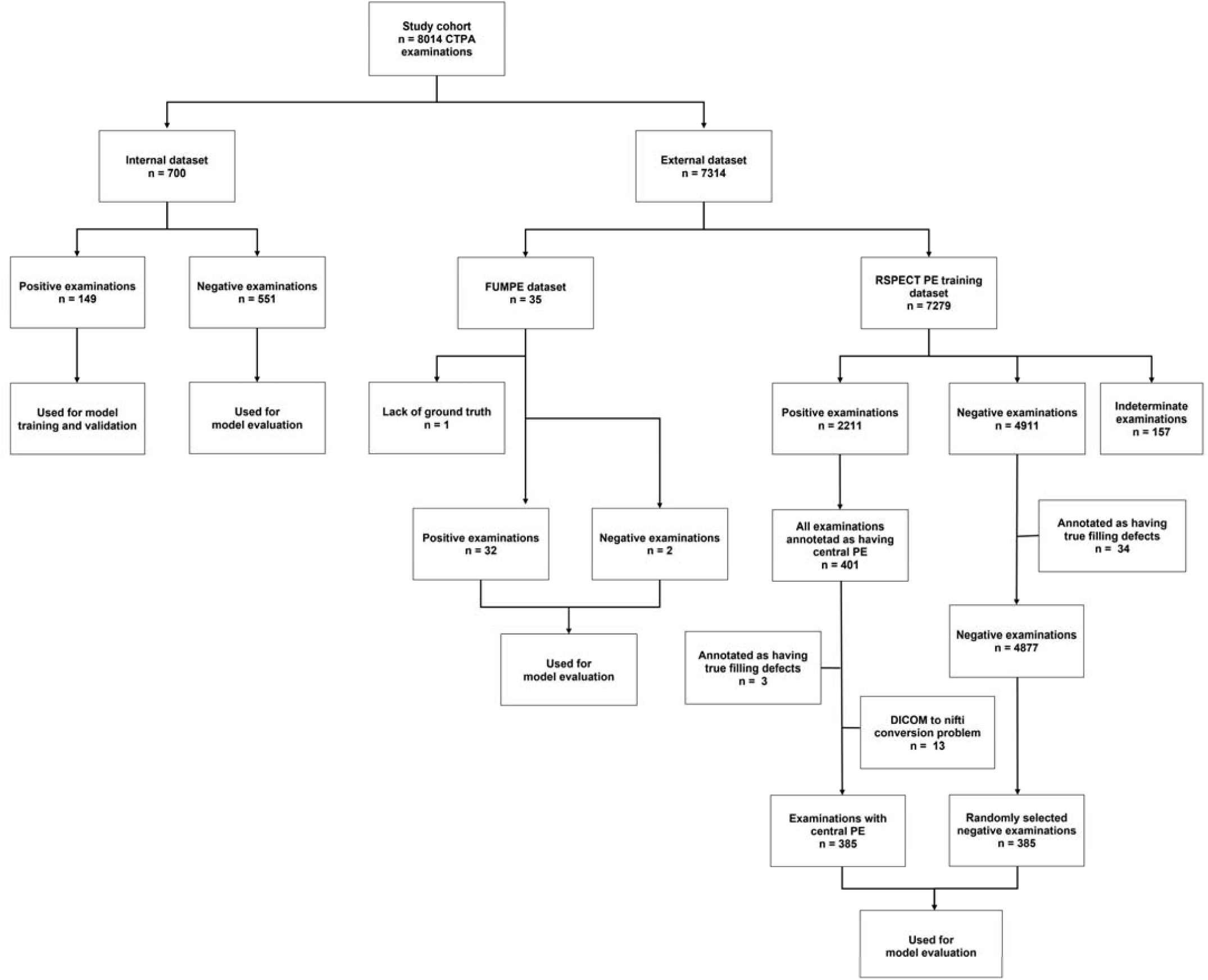
Internal and external datasets for training and evaluation of a segmentation-based classification model for pulmonary embolism detection. Positive examinations refer to the patient having pulmonary embolism (PE) and negative examinations are patients without PE. True filling defect refers to tumor invasion, stump thrombus, catheter, embolized wire, or other obvious non-PE condition as defined in the RSPECT dataset.

### 2.3. nnU-Net Model training and validation

For model training, the nnU-net DL open-source framework, implemented in a Docker container (Docker Inc., Palo Alto, California, USA), was used [19]. The nnU-Net is a semantic segmentation method, and when provided with a training dataset, it automatically configures an end-to-end experimental pipeline (Supp. materials). The PE positive examinations from the internal dataset (n=149) were randomly assigned to training (80%, n=119) and validation sets (20%, n=30) using 5-fold cross-validation during model training (Supp. materials).

### 2.4. Automated Classification Algorithm

After model training and validation, the validated model was embedded in a classification algorithm consisting of three steps, pre-processing, image segmentation inference, and post-processing (Figure 2). Notably, nnU-Net necessitates the utilization of the Neuroimaging Informatics Technology Initiative (NIfTI) file format for model inference. Thus, in the pre-processing step, all DICOM data were converted to the NIfTI format using an in-house Python script. Next, the nnU-Net model inference was performed. Since the nnU-Net model is a volumetric segmentation model, its inference yields a segmentation mask that predicts pulmonary emboli. The segmentation output was transformed into patient-level classification during the post-processing step by applying a threshold to the predicted segmentations, which was based on the total predicted emboli volume. Consequently, a patient-level classification distinguishing between PE and non-PE cases was achieved.

**Figure 2.**
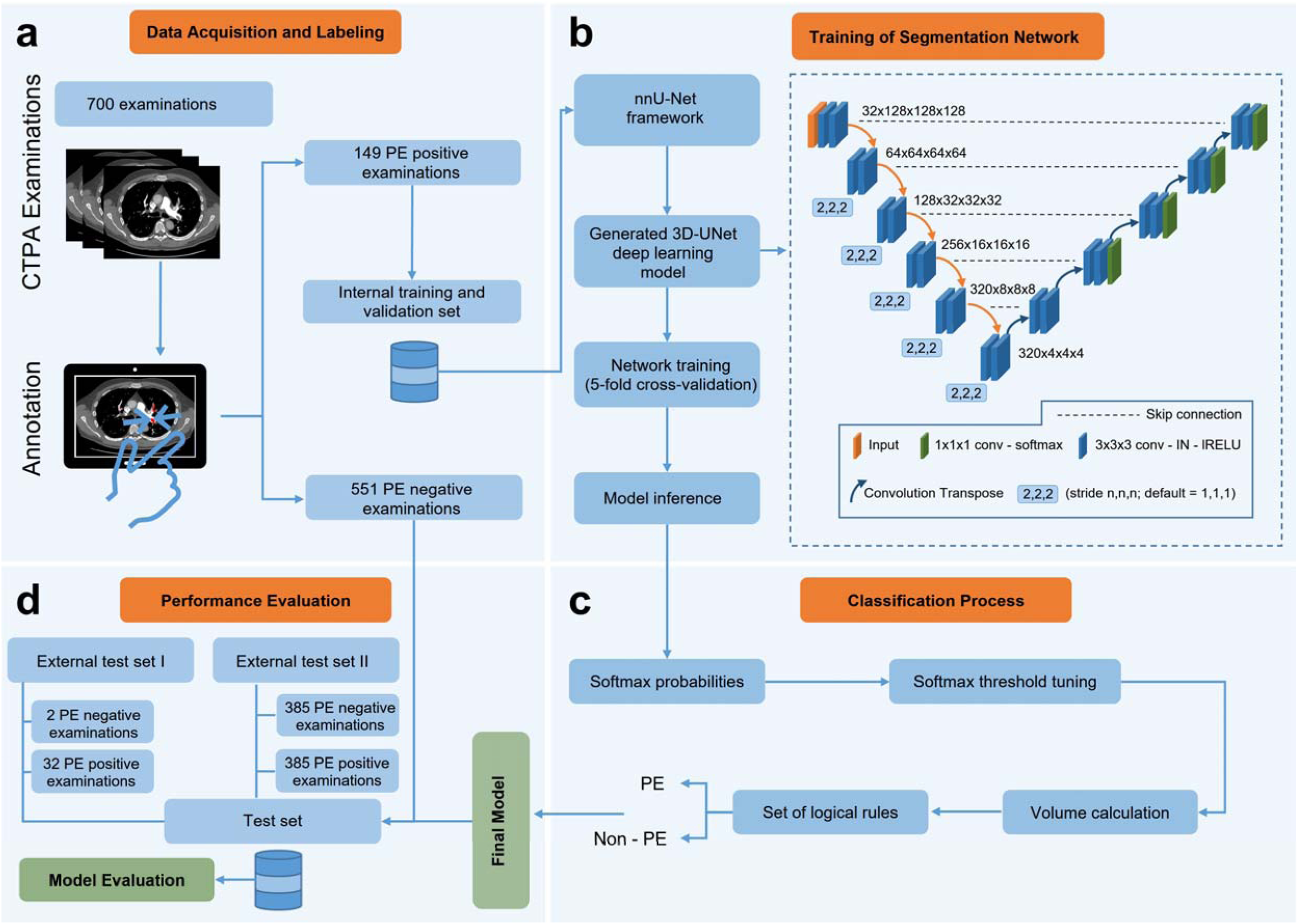
Training and evaluation of a segmentation-based classification model for pulmonary embolism detection. 700 CTPA examinations were collected and annotated by either of two radiologists. Of these, all 149 PE positive examinations were used for training and the PE negative were kept for later evaluation (a). The 3D U-Net deep learning model, which is generated by the nnU-Net framework, was trained with the 149 PE-positive CTPAs using 5-fold cross-validation. The convolution layer used a 3×3×3 filter size by default, followed by an instance normalization (IN) layer and a leaky rectified Linear Unit (lRELU) layer (b). The *softmax* probabilities were obtained from the model inference for fine-tuning classification into PE or non-PE voxel classes and for calculating the predicted PE volume. By thresholding the predicted volumes and applying a set of logical rules, accurate patient-level classification for PE was achieved (c). The final model was evaluated on 804 external CTPA examinations from two publicly available datasets (d).

The softmax activation function in the final layer of the U-Net-like architecture provides a probability distribution across predicted classes. By strategically selecting different softmax probability thresholds, it was possible to generate segmentation masks with varying volumes. As such, we established rules that incorporated different softmax probability thresholds (ranging from 0.75 to 0.95 in 0.05 intervals) and volumetric thresholds (ranging from 0 mm³ to 200 mm³ in 10 mm³ intervals). This approach was instrumental in improving the model’s performance and fine-tuning the differentiation between PE and non-PE voxel classes. Considering these rules, we formulated two distinct strategies. The strategy that offered the best trade-off between sensitivity and specificity was denoted as Strategy 1, while the one delivering the highest specificity was denoted as Strategy 2 (Supp. materials).

### 2.5. Statistical Analysis

Sensitivity and specificity of our trained model for binary classification for PE/non-PE were assessed on a per-patient basis. Matthew’s correlation coefficient (MCC) was used to find the optimal balance between sensitivity and specificity. The area under the receiver operating characteristic (AUROC) curve was used to determine classification performance during model training, validation, and evaluation. Statistical analysis was performed with Microsoft Office Excel (Microsoft Corporation, Washington, USA, Office Professional Plus 2016) and statsmodels package (version 0.13.5) in Python (version 3.8.10; Python Software Foundation). A *p*-value less than .05 was defined as statistically significant and for C.I., the Wilson score interval was used.

## 3. Results

### 3.1. Model training and performance evaluation on the internal dataset

For model training, 2,439,000 voxels of 1497 PE were annotated by two radiologists in all 149 PE positive CTPAs of the internal dataset (Table 1). Acute as well as chronic PEs were annotated, and no distinction was made between them. Consequently, the model did not distinguish between the two types. An nnU-net model was trained with 5-fold cross-validation with 119 training and 30 validation CTPAs per set in 4 sets and 120 training and 29 validation CTPAs in the fifth set without data overlap between the validation sets. To assess model performance, 21 PE positive exams with small PEs (M = 22.9 mm³, SD = 11.6 mm³) with a total volume of less than 50 mm³ were excluded. The remaining 128 PE positive CTPAs and 551 PE negative CTPAs constituted the internal cross-validation and test set. Training and validation was performed on a single Nvidia RTX 2080 TI GPU card which took ∼1 week in total for all cross-validation folds. The classification performance of the trained nnU-Net model on internal and external test datasets was explored over different threshold volumes, with and without post-processing strategies. Without the post-processing strategy and by setting the threshold volume to 20 mm³, a Matthews correlation coefficient score (MCC) of 63.9% was achieved, correctly classifying 128 out of 128 positive examinations as having PE and 433 out of 551 negative examinations as non-PE. With the post-processing strategy 1 (Supp. materials) and threshold volume of 20 mm³, the best MCC (84.9%) was obtained with 123 of 128 positive examinations correctly classified as PE, and 521 of 551 negative examinations correctly classified as non-PE. Further, the model achieved an AUROC of 96.4% and 94.9% with and without post-processing respectively (Figure 3). The trained nnU-Net model thus achieved a sensitivity of 96.1% (95% C.I. 91-98%, *P* < .05) and 100% (95% C.I. 97-100%, *P* < .05), and a specificity of 94.6% (95% C.I. 92-96%, *P* < .05) and 78.6% (95% C.I. 75-82%, *P* < .05) in the internal dataset with and without the post-processing strategies, respectively (Table 2).

**Figure 3.**
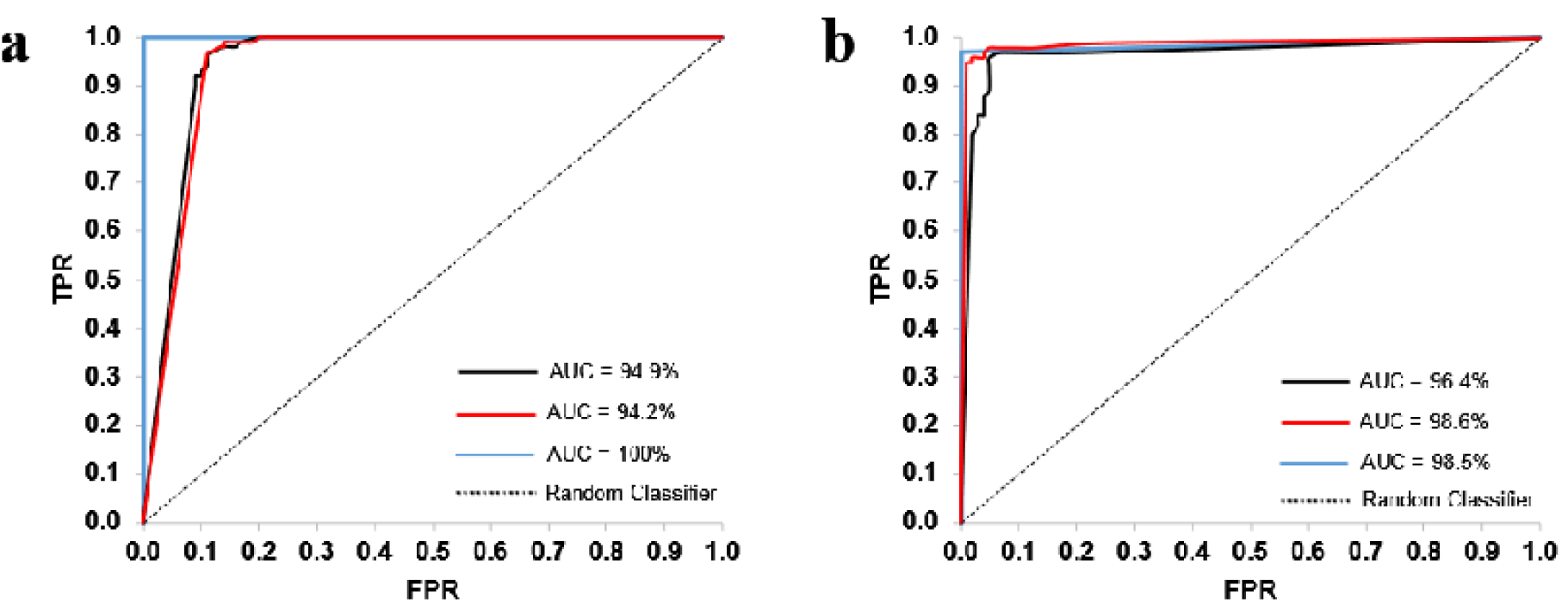
Classification performance of the trained nnU-Net model. Area Under the Curves (AUC) without (a) and with (b) post-processing. Black, internal dataset (n = 679, 128 PE and 551 non-PE); Blue, the FUMPE datasets (n = 34, 32 PE and 2 non-PE); Red, the RSNA PE dataset (n = 770, 385 PE and 385 non-PE). TPR, true positive rate; FPR, false positive rate. AUC values are in percentages.

**Table 1.** Ground truth annotation of 149 internal CTPAs with PE.

| Component | Blood Clots | Average Volume (mm<sup>3</sup>) | Min Volume (mm<sup>3</sup>) | Max Volume (mm<sup>3</sup>) |
| --- | --- | --- | --- | --- |
| 3D (Volume) | 1497 | 682 | 0.21 | 36510 |
| 2D (Area) | 36471 | 16 | 0.21 | 830 |
| 1D (Voxel) | 2439400 |  |  |  |

Note. — The total PE volume in all examinations was 744783 mm<sup>3</sup>. PE = Pulmonary embolism, 3D = 3-dimensional, 2D = 2-dimensional, 1D = 1-dimensional. Min = minimum, Max = maximum. 3D components are composed of 2D components, and 2D components are made up of 1D components. A 1D component, in this context, corresponds to a single voxel.

**Table 2.** Diagnostic performance of the trained model.

| Parameter | Without the Post-Processing |  |  | With the Post-Processing |  |  |
| --- | --- | --- | --- | --- | --- | --- |
|  | Internal Dataset | FUMPE External Dataset | RSNA External Dataset | Internal Dataset | FUMPE External Dataset | RSNA External Dataset |
| No. of CTPAs | 679 | 34 | 770 | 679 | 34 | 770 |
| No. of TN | 433 | 2 | 269 | 521 | 2 | 346 |
| No. of FP | 118 | 0 | 116 | 30 | 0 | 39 |
| No. of TP | 128 | 32 | 385 | 123 | 31 | 379 |
| No. of FN | 0 | 0 | 0 | 5 | 1 | 6 |
| MCC (%) | 63.9 | 100 | 73.3 | 84.9 | 80.4 | 88.6 |
| Sensitivity (%) | 100 (97-100) | 100 (89-100) | 100 (99-100) | 96.1 (91-98) | 96.9 (84-99) | 98.4 (97-99) |
| Specificity (%) | 78.6 (75-82) | 100 (34-100) | 69.9 (65-74) | 94.6 (92-96) | 100 (34-100) | 89.9 (86-93) |
| Accuracy (%) | 82.6 | 100 | 84.9 | 94.8 | 97.1 | 94.2 |
| Balanced Accuracy (%) | 89.3 | 100 | 84.9 | 95.4 | 98.4 | 94.2 |
| AUC (%) | 94.9 | 100 | 94.2 | 96.4 | 98.5 | 98.6 |

Note. — The threshold volume is set to 20 mm<sup>3</sup>. Data in parentheses are 95% CIs in percentages. CTPAs = computed tomography (CT) pulmonary angiography (CTPA) examinations, TN = true-negative CTPAs, FP = false-positive CTPAs, TP = true-positive CTPAs, FN = false-negative CTPAs, MCC = Matthew's correlation coefficient, AUC = area under the receiver operating characteristic curve.

### 3.2. Model performance on external datasets

For external evaluation, the trained model was applied to a total of 804 CTPAs from two publicly available datasets. First, 34 PE positive CTPAs and 2 PE negative CTPAs from the FUMPE dataset were analyzed. With post-processing strategy 1, an MCC score of 80.4% was obtained with 31 of 32 positive examinations correctly classified as PE, and 2 of 2 negative examinations correctly classified as non-PE. The trained model achieved AUROC 98.5% (Figure 3) with sensitivity of 96.9% (95% C.I. 84-99%, *P* < .05) and specificity of 100% (95% C.I. 34-100%, *P* < .05) (Table 2, Supp. Table 5). Focusing on central PE, where the annotations can be assumed to be more consistent, we used 385 CTPAs annotated as having at least one central PE and 385 PE negative CTPAs from the RSPECT pulmonary embolism CT dataset for model evaluation. With the post-processing strategy 1 (Supp. Materials), an MCC of 88.6% was obtained with 379 of 385 positive examinations correctly classified as PE, and 346 of 385 negative examinations correctly classified as non-PE. The trained model achieved an AUROC of 98.6% (Figure 3) with sensitivity of 98.4% (95% C.I. 97-99%, *P* < .05) and a specificity of 89.9% (95% C.I. 86-93%, *P* < .05) (Table 2, Supp. Table 6). Without the post-processing strategy and by setting the threshold volume to 20 mm³, MCC of 100% and 73.3% were obtained with 32 (n=32) and 385 (n=385) positive examinations correctly classified as PE, and 2 (n=2) and 269 (n= 385) negative examinations correctly classified as non-PE in the first and second external datasets, respectively (Table 2, Supp. Table 2 and 3). Moreover, the model achieved an AUROC of 100% and 94.2% (Figure 3) with a sensitivity of 100% (95% C.I. 89-100%, *P* < .05) and 100% (95% C.I. 99-100%, *P* < .05), and a specificity of 100% (95% C.I. 34-100%, *P* < .05) and 69.9% (95% C.I. 65-74%, *P* < .05) in the first and second datasets, respectively (Table 2, Supp. Table 2 and 3).

The output of the automated classification algorithm is shown (Figure 4, Supp. Figures 1-3). Performing model inference within the nnU-Net framework for a single CTPA volume examination, utilizing a singular Nvidia RTX 4090 GPU card, required 240-300 s. Furthermore, disabling the test time augmentation (TTA) yielded a decrease in inference time to 60-70 s with an average accuracy decline of 1% (Supp. Tables 1-12).

**Figure 4.**
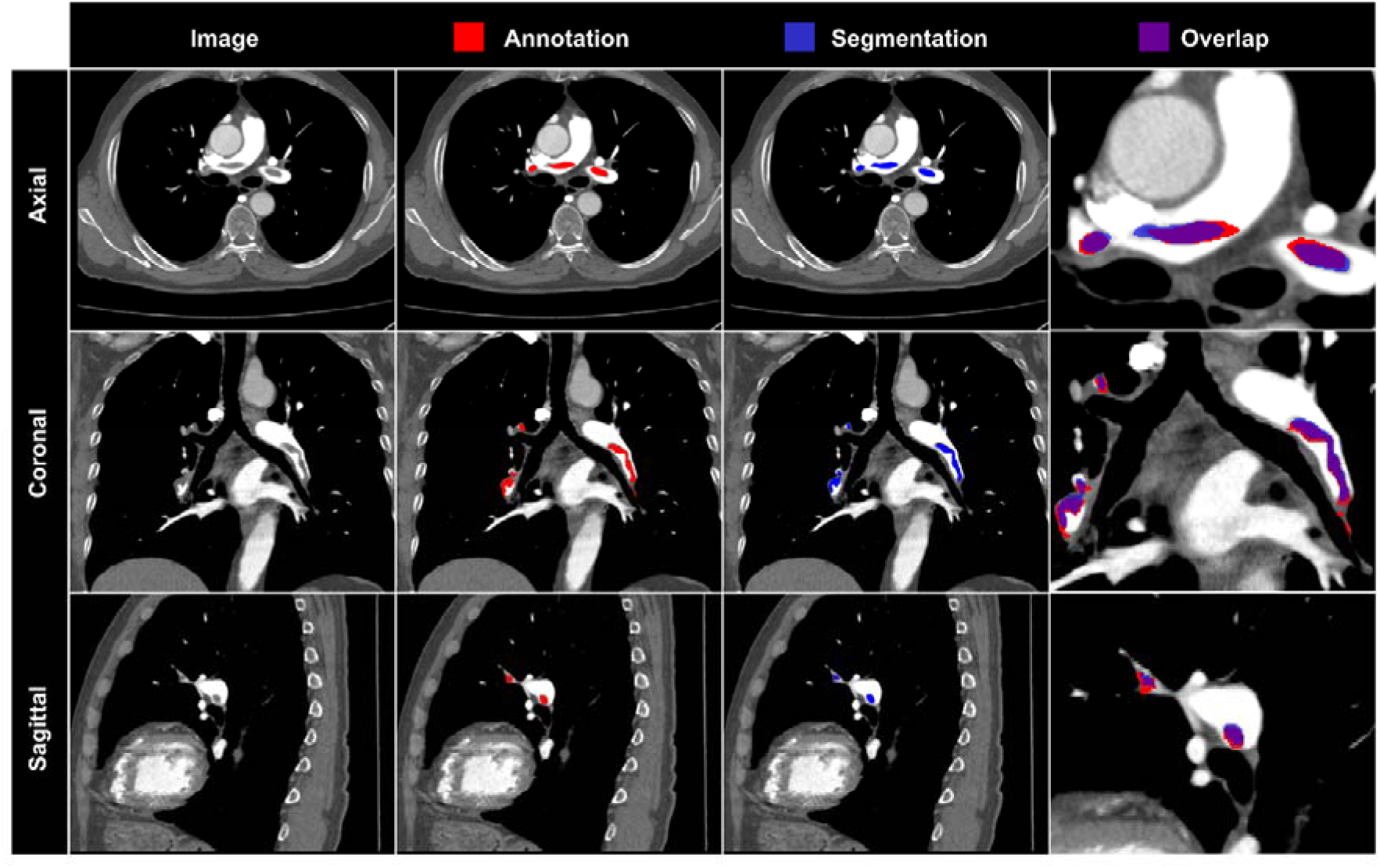
Representative segmentation results of the trained model. Axial, coronal, and sagittal planes from the same CTPA examinations from the external FUMPE dataset with the same window setting (width = 800 HU, level = 100 HU) are shown. Red, pulmonary embolism annotation; blue, model segmentation; purple, overlay of annotation and model segmentation.

### 3.3. Benchmarking of model performance

As mentioned above, post-processing strategy 1 was used to find out the best balance between sensitivity and specificity and 20 mm³ was determined as the optimal threshold volume. Aiming for the highest specificity and the lowest patient level false positive rate, we used post-processing strategy 2 where 50 mm³ was determined as optimal threshold volume. For size comparison, 20 mm³, 50 mm³, and other threshold volumes (Figure 5a) are compared to a segmented reference pulmonary artery (Figure 5b). In the internal test dataset, the highest specificity (96.7%; 95% C.I. 95-98%, *P* < .05) was obtained with a sensitivity of 87.5% (95% C.I. 81-92%, *P* < .05) with post-processing strategy 2 (Supp. materials, Supp. Table 7) and threshold volume of 50 mm³. For the external datasets, the highest specificity 100% (95% C.I. 34-100%, *P* < .05) and 96.9% (95% C.I. 95-98%, *P* < .05) was obtained with a sensitivity of 90.6% (95% C.I. 76-97%, *P* < .05) and 96.6% (95% C.I. 94-98%, *P* < .05) in the FUMPE and RSPECT datasets, respectively (Supp. Table 8 and 9). Moreover, we examined the source of false positives by implementing post-processing strategy 2, which led to a minimum number of FPs per dataset. The most frequent false positives were due to low contrast medium in pulmonary arteries (Table 3). Whereas 18% of FPs occurred on the outside of the thoracic cavity, in the upper abdomen, or in the superior vena cava, the remaining FPs occurred within or close to the pulmonary vessel network. Besides, most of the false negatives (FN) occurred in the RSPECT dataset, and the primary cause of these FNs is chronic PEs. We next compared model performance to those of previous studies (Table 4). With post-processing strategy 2, the proposed algorithm achieved a sensitivity of 96.2% (95% C.I. 94-98%, *P* < .05) and a specificity of 96.8% (95% C.I. 95-98%, *P* < .05) on the combined (internal and external) testing set (Supp. Table 12). While investigating the causes of false positives, we observed that 3 CTPAs from the RSPECT dataset that were annotated as PE negative were actually PE positive. Considering this correction, the proposed algorithm achieved a specificity of 97.1%.

**Figure 5.**
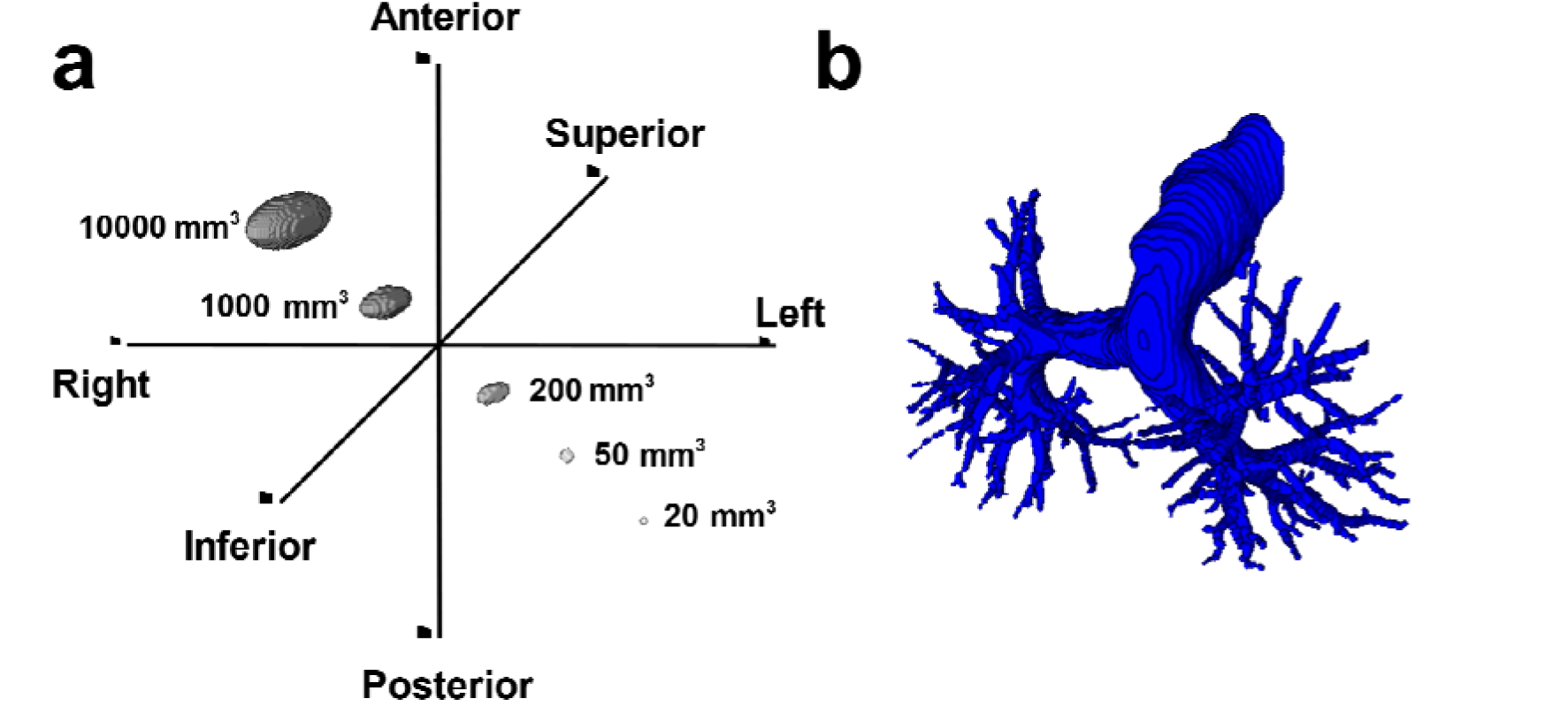
An illustration of a segmented reference pulmonary artery with reference threshold volumes. Patient orientation of 3D volumes with reference threshold volumes (20, 50, 200, 1000, and 10000 mm³) (a). Manually segmented reference pulmonary artery (volume of 113 cm³) from a male patient without PE (b). All volumetric images are isotropic (1 mm × 1 mm × 1 mm).

**Table 3.** Sources of false positives in PE negative CTPA examinations from internal and external datasets.

| Source | Internal Dataset<br>(n=18) | RSPECT External Dataset<br>(n=12) |
| --- | --- | --- |
| Flow artifact | 1 | 1 |
| Upper abdomen (in left colon) | 1 | 0 |
| Outside the thoracic cavity | 3 | 0 |
| Low contrast medium in PT | 6 | 2 |
| Pulmonary vein | 1 | 2 |
| Superior vena cava | 1 | 0 |
| Intrafissural fluid / atelectasis | 0 | 1 |
| Multiple metastasis | 1 | 1 |
| Tumor | 4 | 2 |
| True pulmonary emboli | 0 | 3 |

Note. — CTPAs = computed tomography (CT) pulmonary angiography (CTPA) examinations, PE = pulmonary embolism, PT = pulmonary trunk, RSPECT = RSNA Pulmonary Embolism CT Dataset. The cause of false positives in a total of 30 CTPAs is shown, 18 in internal and 12 in external datasets.

**Table 4.**
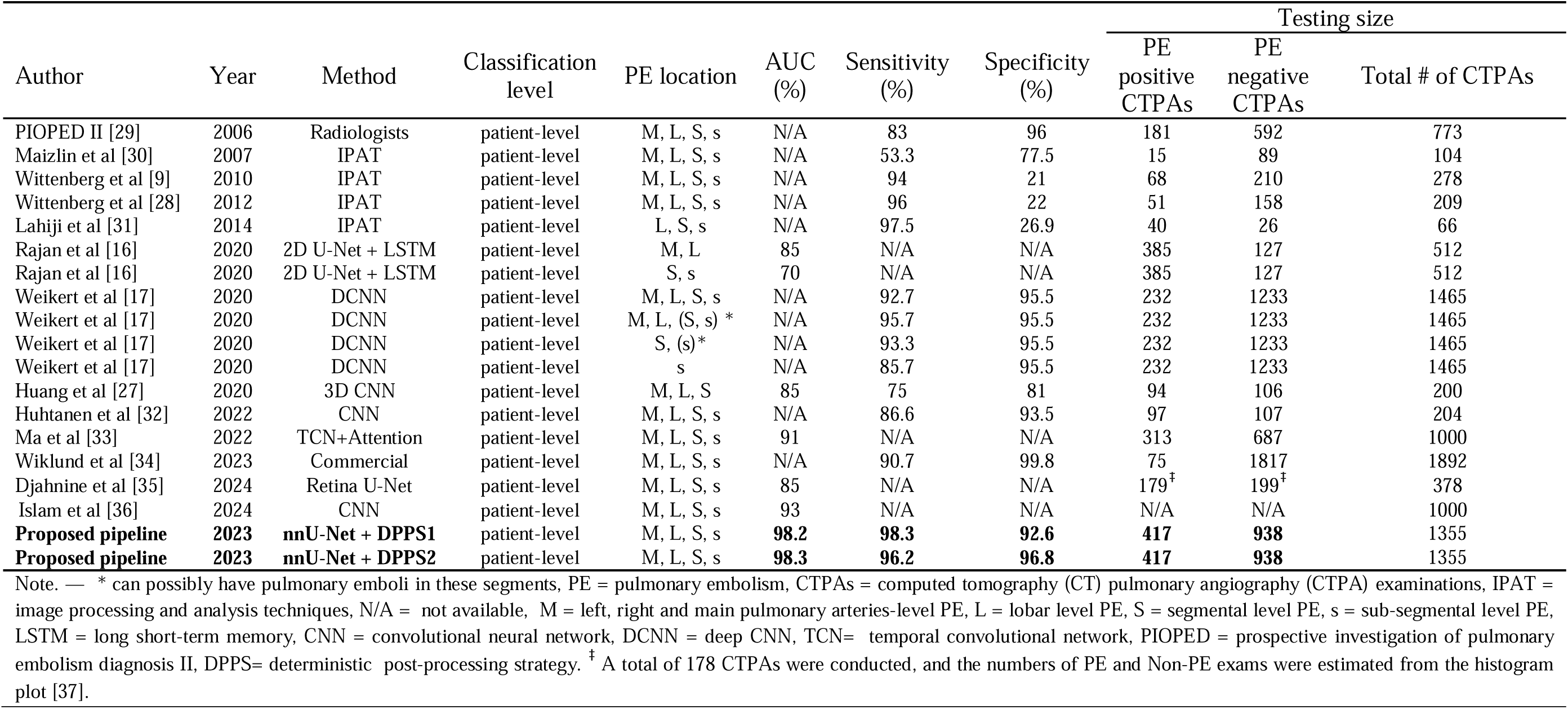
Model performance comparison for patient-level classification for PE in CTPA examinations.

| Author | Year | Method | Classification level | PE location | AUC (%) | Sensitivity (%) | Specificity (%) | Testing size |  |  |
| --- | --- | --- | --- | --- | --- | --- | --- | --- | --- | --- |
|  |  |  |  |  |  |  |  | PE positive CTPAs | PE negative CTPAs | Total # of CTPAs |
| PIOPED II [29] | 2006 | Radiologists | patient-level | M, L, S, s | N/A | 83 | 96 | 181 | 592 | 773 |
| Maizlin et al [30] | 2007 | IPAT | patient-level | M, L, S, s | N/A | 53.3 | 77.5 | 15 | 89 | 104 |
| Wittenberg et al [9] | 2010 | IPAT | patient-level | M, L, S, s | N/A | 94 | 21 | 68 | 210 | 278 |
| Wittenberg et al [28] | 2012 | IPAT | patient-level | M, L, S, s | N/A | 96 | 22 | 51 | 158 | 209 |
| Lahiji et al [31] | 2014 | IPAT | patient-level | L, S, s | N/A | 97.5 | 26.9 | 40 | 26 | 66 |
| Rajan et al [16] | 2020 | 2D U-Net + LSTM | patient-level | M, L | 85 | N/A | N/A | 385 | 127 | 512 |
| Rajan et al [16] | 2020 | 2D U-Net + LSTM | patient-level | S, s | 70 | N/A | N/A | 385 | 127 | 512 |
| Weikert et al [17] | 2020 | DCNN | patient-level | M, L, S, s | N/A | 92.7 | 95.5 | 232 | 1233 | 1465 |
| Weikert et al [17] | 2020 | DCNN | patient-level | M, L, (S, s) * | N/A | 95.7 | 95.5 | 232 | 1233 | 1465 |
| Weikert et al [17] | 2020 | DCNN | patient-level | S, (s)* | N/A | 93.3 | 95.5 | 232 | 1233 | 1465 |
| Weikert et al [17] | 2020 | DCNN | patient-level | s | N/A | 85.7 | 95.5 | 232 | 1233 | 1465 |
| Huang et al [27] | 2020 | 3D CNN | patient-level | M, L, S | 85 | 75 | 81 | 94 | 106 | 200 |
| Huhtanen et al [32] | 2022 | CNN | patient-level | M, L, S, s | N/A | 86.6 | 93.5 | 97 | 107 | 204 |
| Ma et al [33] | 2022 | TCN+Attention | patient-level | M, L, S, s | 91 | N/A | N/A | 313 | 687 | 1000 |
| Wiklund et al [34] | 2023 | Commercial | patient-level | M, L, S, s | N/A | 90.7 | 99.8 | 75 | 1817 | 1892 |
| Djahnine et al [35] | 2024 | Retina U-Net | patient-level | M, L, S, s | 85 | N/A | N/A | 179<sup>‡</sup> | 199<sup>‡</sup> | 378 |
| Islam et al [36] | 2024 | CNN | patient-level | M, L, S, s | 93 | N/A | N/A | N/A | N/A | 1000 |
| **Proposed pipeline** | **2023** | **nnU-Net + DPPS1** | patient-level | M, L, S, s | **98.2** | **98.3** | **92.6** | **417** | **938** | 1355 |
| **Proposed pipeline** | **2023** | **nnU-Net + DPPS2** | patient-level | M, L, S, s | **98.3** | **96.2** | **96.8** | **417** | **938** | 1355 |

Note. — \* can possibly have pulmonary emboli in these segments, PE = pulmonary embolism, CTPAs = computed tomography (CT) pulmonary angiography (CTPA) examinations, IPAT = image processing and analysis techniques, N/A = not available, M = left, right and main pulmonary arteries-level PE, L = lobar level PE, S = segmental level PE, s = sub-segmental level PE, LSTM = long short-term memory, CNN = convolutional neural network, DCNN = deep CNN, TCN= temporal convolutional network, PIOPED = prospective investigation of pulmonary embolism diagnosis II, DPPS= deterministic post-processing strategy. <sup>‡</sup> A total of 178 CTPAs were conducted, and the numbers of PE and Non-PE exams were estimated from the histogram plot [37].

## 4. Discussion

Detection of PE in CTPA using deep convolutional neural networks (DCNN) was first demonstrated by Tajbakhsh et al. with a sensitivity of 83% and 34.6% at 2 FPs per examinations on 121 internal and 20 external test examinations, respectively [26]. Rajan et al. proposed a two-stage solution where a 2D U-Net model was used for PE candidate generation, followed by a convolutional long short-term memory (LSTM) network coupled with multiple instance learning to detect PE lesions, with AUROC of 70% for subsegmental and segmental PE and 0.85 for saddle and main pulmonary artery PE on a test dataset of 512 CTPA examinations [16]. In a study by Huang et al. [27], a 3D CNN model termed PENet was developed which achieved a sensitivity of 75% and specificity of 81% on an external test dataset of 200 CTPA examinations. However, all these studies have major limitations such as small testing dataset sizes or low specificity rates. The current state-of-the-art results were recently achieved using the Resnet architecture on 1465 CTPA examinations with a sensitivity of 92.7% and specificity of 95.5% at the patient level [18]. Taken together, the performance of AI systems for PE detection is now at a point where clinical utility can be expected, but further gains in sensitivity and specificity are still warranted.

In this study, we developed an algorithm that classifies CTPA examinations for presence of PE consisting of two main stages, PE candidate selection and post-processing. For PE candidate selection, we trained and validated a semantic segmentation model, nnU-Net, on our internal dataset. The nnU-Net is a medical image segmentation framework based on the U-Net architecture and has outperformed state-of-the-art models by competing in 53 segmentation tasks from 11 international biomedical image segmentation challenges and taking first place in 33 of them [19]. To our knowledge, this is the first use of nnU-Net for classification for PE. To transform the segmentation model into a classification model, we developed rules based on probability and minimum volume thresholds as a post-processing stage. We defined two post-processing strategies, one for the best trade-off between sensitivity and specificity and one for achieving the highest specificity. At the best trade-off between sensitivity and specificity, the patient-level classification performance of the trained model achieved a sensitivity of 98.3% and specificity of 92.6% on the combined testing dataset using a threshold volume of 20 mm³, compared to specificity of 75.1% with sensitivity of 100% without post-processing. Thus, by sacrificing 1.7% of sensitivity, the model gained 17.4% in specificity using post-processing. The model outperformed the current state-of-the-art using the strategy of highest specificity, achieving 96.2% sensitivity and a specificity of 96.8% on the combined testing dataset of 1355 CTPA examinations with a total emboli volume threshold of 50 mm³.

Treatment for small PEs is debated and controversial [24,25] and PE volumes in this context are rarely measured and reported in the literature and are subject to inter-observer variability. We decided to set a cut-off volume to exclude the very smallest and in some cases doubtful PEs in our internal dataset. Radiologists DT and TF segmented all 1497 PEs and found a total volume of 50 mm³ to be a reasonable cut-off for exclusion. In a scenario where the total PE volume of 50 mm³ was represented by a single, equidimensional embolus, the cut-off size would amount to a cylinder with a diameter and height of 4 mm. Many of the PEs in the 21 excluded exams were much smaller eccentric or tiny webs, likely chronic.

Although the nnU-Net based model presented here is superior to the state-of-the-art, there are some limitations and opportunities for future enhancement. First, the model was trained on data from a single institution, although derived from five different CT scanners. Second, the training dataset, even though the proportion was not analyzed in detail, was predominantly comprised of acute PEs with a limited representation of chronic PEs. However, our observations suggest that chronic PEs within the RSPECT dataset are a significant factor contributing to false negatives. Third, the RSPECT validation dataset lacks voxel level annotation of PE by radiologists, which precludes final determination of sensitivity and specificity until a review has been completed. Finally, the activation of test time augmentation (TTA) extends the model inference duration to ∼300 s per CTPA examination. Conversely, deactivating the TTA reduces the model inference time to a range of 60 to 70 seconds for a single CTPA examination. However, when TTA is disabled, sensitivity and specificity decrease by approximately 0.3% and 1.7%, respectively.

In conclusion, nnU-Net deep learning based binary classification for PE holds potential to assist radiologists in the reading of CTPA examinations. Preferentially, such a system could prioritize PE positive cases in the work list, identifying high-priority cases for swift review [28], or provide a second opinion.

## Data Availability

Data generated or analyzed during the study are available from the corresponding author by request.

## Data sharing statement

Data generated or analyzed during the study are available from the corresponding author by request.

## Acknowledgements

The project was supported by a grant from Analytic Imaging Diagnostics Arena (AIDA) to Tobias Sjöblom. Tomas Fröding and Dimitrios Toumpanakis were financially supported by clinical fellowships from AIDA. Tomas Fröding was financially supported by the Centre for Clinical Research Sörmland, Uppsala University.

## Supplementary Materials

### S.1. Dataset

#### S.1.1 Internal dataset CT Acquisition Protocols

All 700 CTPAs were performed with bolus tracking technique with the region of interest (ROI) in the pulmonary trunk. Different Hounsfield unit thresholds and delays were used. Contrast medium doses were recorded for 191 (range 20 ml – 114 ml, mean 62 ml) and injection rates for 158 (range 2,4 ml/s – 6,1 ml/s, mean 3,6 ml/s) CTPAs, respectively. The most frequently used CT image acquisition parameters were slice thickness 0.625 mm (range 0.625 mm - 2.0 mm), pixel spacing 0.7 mm (range 0.59 mm - 0.98 mm), tube voltage 100 kV (range 80 kV - 120 kV) and scanning direction caudal to cranial. The CTPAs acquired on the Siemens Somatom Definition Flash CT were in the majority of cases performed with dual-energy source acquisition with tube settings 80 kV / 140 kV and the images used in the dataset were post-processed blended images from a weighting factor 0.5.

#### S.1.2 Distribution of CT Pulmonary Angiography examinations from the same patient in internal datasets

The internal dataset comprises 700 CT Pulmonary Angiography (CTPA) examinations involving 652 patients. Among them, 41 patients underwent CTPA twice, 2 patients thrice, and 1 patient four times. Of the 149 pulmonary embolism (PE) -positive examinations, 142 patients were involved, and of the 551 PE-negative examinations, 520 patients were included. Ten patients had both PE-negative and PE-positive CT examinations. Since the CT scans acquired from the same patient were performed at different occasions, several anatomical aspects depending on breath hold level, angle of spine and pulmonary disease status were different (Supp. Figures 4-5). This means that there were differences at the voxel level and also differences in the data label (positive/negative PE), depending on the scan session. The examinations were therefore used and analyzed as if they had been obtained from different patients. They were therefore randomly distributed to cross-validation dataset, regardless of whether they belonged to the same patient. In the external datasets, all CTPAs were obtained from different patients and thus truly statistically independent.

#### S.1.3 Ferdowsi University of Mashhad’s Pulmonary Embolism Dataset

The Ferdowsi University of Mashhad’s PE dataset (FUMPE) is a publicly available dataset consisting of 35 CTPA examinations with voxel-level PE annotations by radiologists. One PE-positive examination was excluded due to a lack of ground truth annotation. Out of the 34 CTPA examinations, 32 were PE-positive and 2 were PE-negative. When examining the ground truth, we noticed that the slice locations of PE annotations were incorrect in 8 CTPA examinations. Specifically, in these cases, PE annotations that should have been located in slice 101 were mistakenly placed in slice 11. As a result, we relocated the PE annotations from slice 11 to slice 101.

### S.2. Environmental Settings and Versions

#### S.2.1 Model Training Environment

The environmental settings employed for both model training and cross-validation in this study encompass specific software versions: Ubuntu 22.04.3 LTS as the operating system, Docker version 24.0.7 for containerization, and CUDA Version 12.1, along with Nvidia driver version 530.30.02, are employed to facilitate seamless interactions with the NVIDIA GeForce RTX 2080 Ti GPUs. The programming language employed is Python, specifically version 3.8.10. The deep learning framework PyTorch is leveraged in version 2.0.0, and the semantic segmentation method nnU-Net is implemented in version 1.7.1.

#### S.2.2 Model Inference Environment

For model inference, another workstation was utilized, with specific environmental settings and software versions. These settings include Ubuntu 22.04.3 LTS as the operating system, Docker version 24.0.7 for containerization, CUDA Version 12.2, and Nvidia driver version 535.129.03, facilitating seamless interactions with the NVIDIA GeForce RTX 4090 GPU. The programming language employed is Python version 3.10.6. PyTorch, the deep learning framework, is utilized in version 2.1.0, and the semantic segmentation method nnU-Net is implemented in version 1.7.1.

### S.3. The nnU-Net Deep Learning Framework

#### S.3.1 Hyperparameters

In the training of our deep learning model, the nnU-Net framework employed a specific set of hyperparameters to optimize the learning process. The chosen optimizer is Stochastic Gradient Descent (SGD) with Nesterov momentum, utilizing a momentum value of 0.99. Additionally, weight decay was incorporated with a coefficient of 3e-05 to regulate the model’s complexity during training. The initial learning rate was set to 0.01, providing a starting point for the optimization process. To enhance the training procedure, a learning rate scheduler was implemented with a patience parameter of 30 epochs and a threshold of 1e-06. Lastly, the maximum number of training epochs was defined as 1000. These carefully selected hyperparameters contribute to the fine-tuning of the model, optimizing its performance over the course of training.

#### S.3.2 Data Augmentation

The nnU-Net framework employed a set of data augmentation techniques to generalize the models to prevent overfitting to the training data set. Elastic deformation, a spatial transformation technique, was introduced with an alpha range of (0.0, 200.0) and a sigma range of (9.0, 13.0), implemented with a probability of occurrence set at 0.2. Scaling transformations were applied within the range of (0.7, 1.4). To introduce variability in the orientation of the input data, rotational transformations along the X, Y, and Z axes were implemented with specified ranges. Gamma correction, an intensity transformation, was applied with a probability of 0.3 and a gamma range of (0.7, 1.5). Mirroring along axes (0, 1, 2) were applied. Furthermore, a cascaded random binary transformations and additive brightness adjustments were employed with specified probabilities and parameters.

#### S.3.3 3D U-Net Architecture

The nnU-Net framework is configured to generate a 3D U-Net architecture for semantic segmentation tasks. The 3D U-Net architecture is characterized by a symmetrical design with a decoder path that uses transposed convolutions for up sampling. The decoder path of the network consists of five transposed convolutional layers. Each layer employs 3D transposed convolutional operation with varying input and output channel sizes, effectively increasing spatial resolution. Starting with the first layer, it utilizes a transposed convolution operation with 320 input channels, 320 output channels, a 2x2x2 kernel size, and a stride of 2 in all spatial dimensions. Subsequent other layers follow a similar structure, progressively decreasing the number of input channels while maintaining the up-sampling strategy. Additionally, the architecture includes an encoder path with five convolutional layers, where each 3D convolutional operation employs a 1x1x1 kernel with a stride of 1. These layers reduce the channel depth and capture hierarchical features. Each convolutional layer followed by 3D instance normalization and leaky rectified linear unit activation.

## S.4. Post-processing step

The nn-Unet *softmax* activation function of the final layer of the U-Net architecture can be used to scale network output into probabilities. Hence, the probabilities could be gathered, and not only final pixel class values. We developed a set of logical rules based on different *softmax* probability thresholds (0.75 - 0.95) and threshold volumes per examinations (0 mm³ to 200 mm³ in 10 mm³ intervals) to reduce false positives (FPs) and convert nnU-Net inference segmentation output into a patient-level classification output. By setting different *softmax* probability thresholds, we obtained different predicted PE volumes. If the model is well-trained to distinguish between PE and non-PE classes, the number of predicted voxels (false positive voxels) that do not belong to the PE class will decrease when the *softmax* probabilities are set to higher thresholds. Therefore, we developed the formulas below to decide whether the total predicted PE volume is sufficient to determine the patient as PE positive/negative.

Proposition 1:

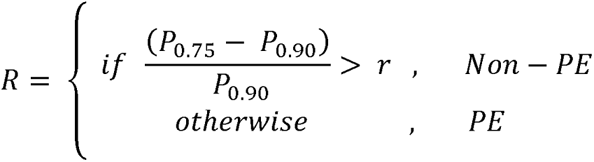

where *P*_0.75_ is the volume of total PE predicted by the trained model at a softmax probability of 0.75, *P*_0.90_ the ratio factor, which was fixed at 15. The *softmax* probability value range and the ratio factor were optimized by systematic exploration.

Proposition 2:

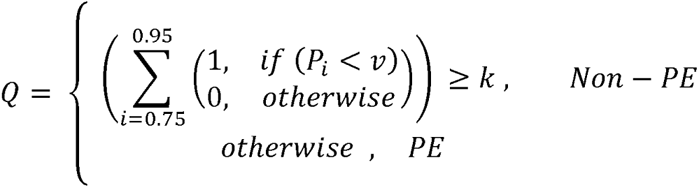

where *P_i_* is the volume of total PE predicted by the trained model at a *softmax* probability of *i* between 0.75 to 0.95 with 0.05 intervals, *v* is the threshold volume between 0 and 200 mm³ at 10 mm³ intervals and is the condition factor (min value is 0, max value is 4) that refers to the total number of true conditions satisfying *P_i <_ v* equation.

Then, the final decision is made as follows:

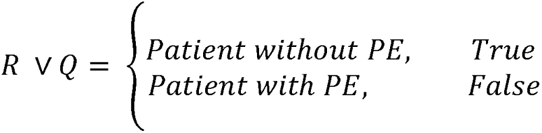

According to the propositions above, we defined two post-processing strategies. Strategy 1 (Rule-in classification for PE) aimed to find the exact threshold volume value and value for the best trade-off between sensitivity and specificity by checking the Matthew’s correlation coefficient (MCC) value. And strategy 2 (Rule-out classification for PE) aimed to find the exact threshold volume and values for the highest specificity alongside the highest MCC value.

*Strategy 1:*

By systematic exploration, setting the threshold volume value to 20 mm³ and the k value to 1 gives the highest MCC value (84.9%, Supplementary Table 4).

*Strategy 2*

By systematic exploration, setting the threshold volume value to 50 mm³ and the k value to 0 gives the highest specificity alongside the highest MCC value (83.7%, Supplementary Table 7).

## Abbreviations

PE: pulmonary embolism
CTPA: computed tomography pulmonary angiography
nnU-Net: no-new-U-Net
DL: deep learning
CADe: computer-aided detection.

---

**Supplemental Figure 1.**
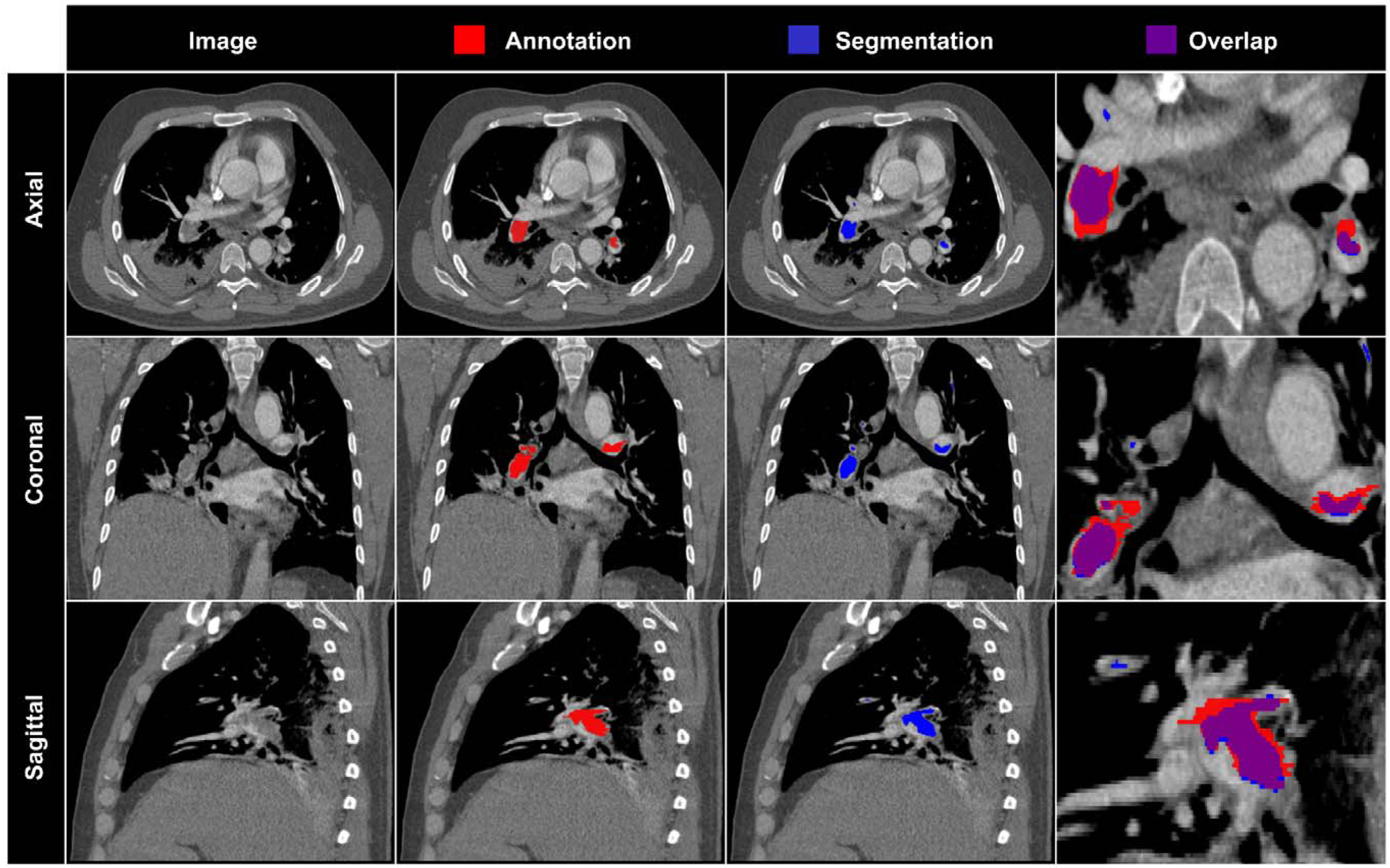
Representative segmentation results from the FUMPE dataset (patient 03). Axial, coronal, and sagittal planes from the same CTPA examination from the external FUMPE dataset with the same window setting (width = 800 HU, level = 100 HU) are shown. Red, pulmonary embolism annotation; blue, model segmentation; purple, overlay of annotation and model segmentation.

**Supplemental Figure 2.**
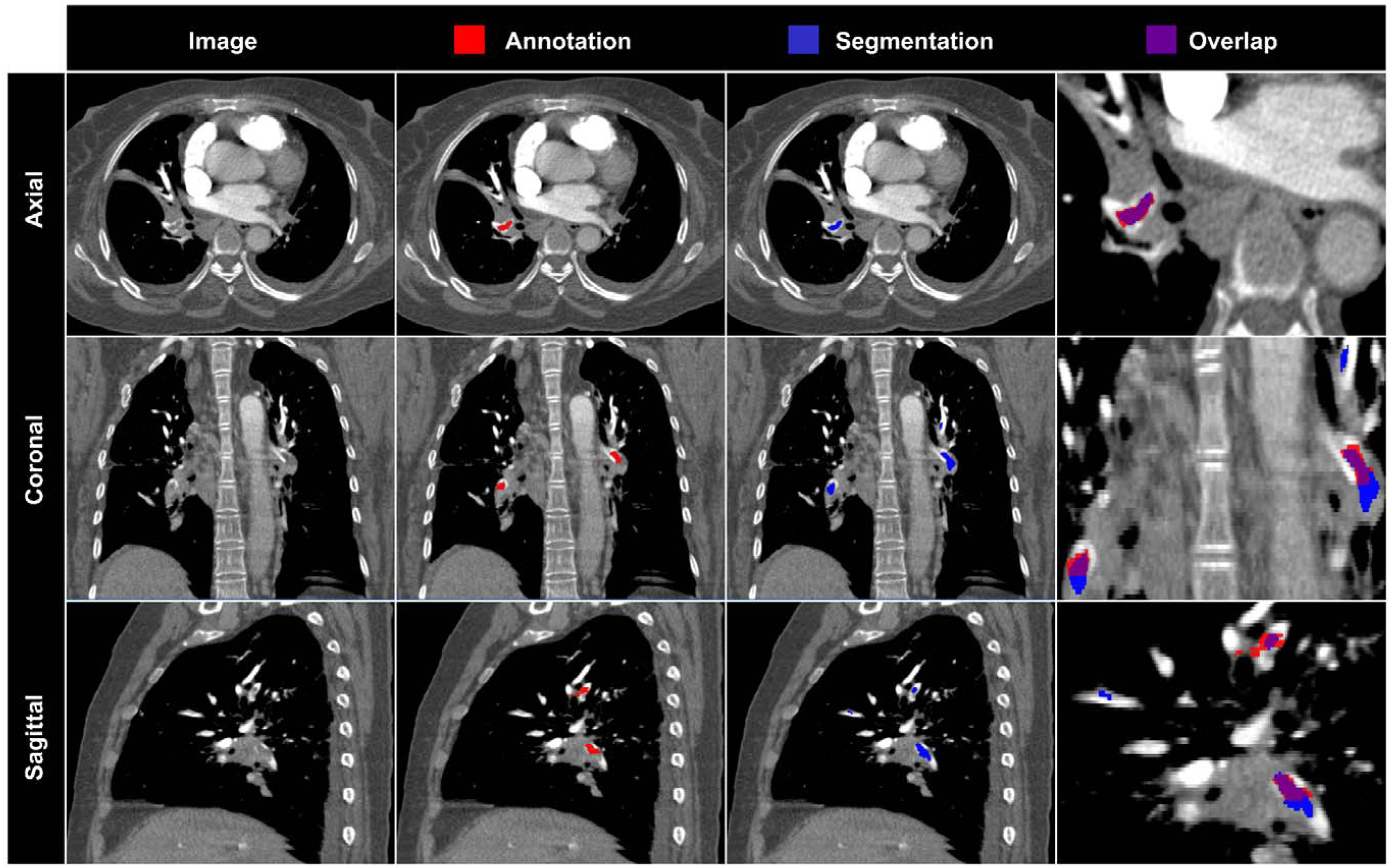
Representative segmentation results from the FUMPE dataset (patient 04). Axial, coronal, and sagittal planes from the same CTPA examination from the external FUMPE dataset with the same window setting (width = 800 HU, level = 100 HU) are shown. Red, pulmonary embolism annotation; blue, model segmentation; purple, overlay of annotation and model segmentation.

**Supplemental Figure 3.**
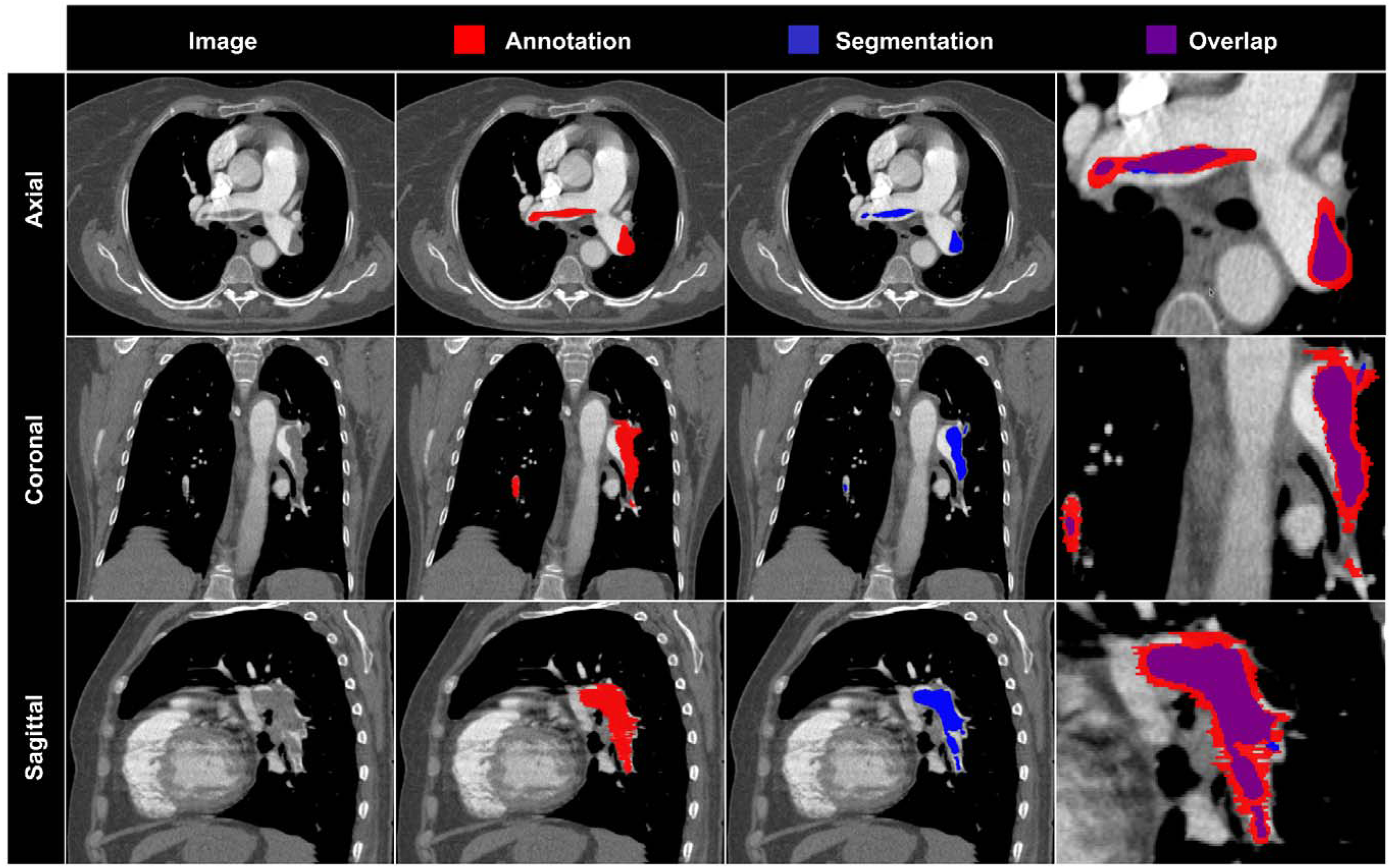
Representative segmentation results from the FUMPE dataset (patient 05). Axial, coronal, and sagittal planes from the same CTPA examination from the external FUMPE dataset with the same window setting (width = 800 HU, level = 100 HU) are shown. Red, pulmonary embolism annotation; blue, model segmentation; purple, overlay of annotation and model segmentation.

**Supplemental Figure 4.**
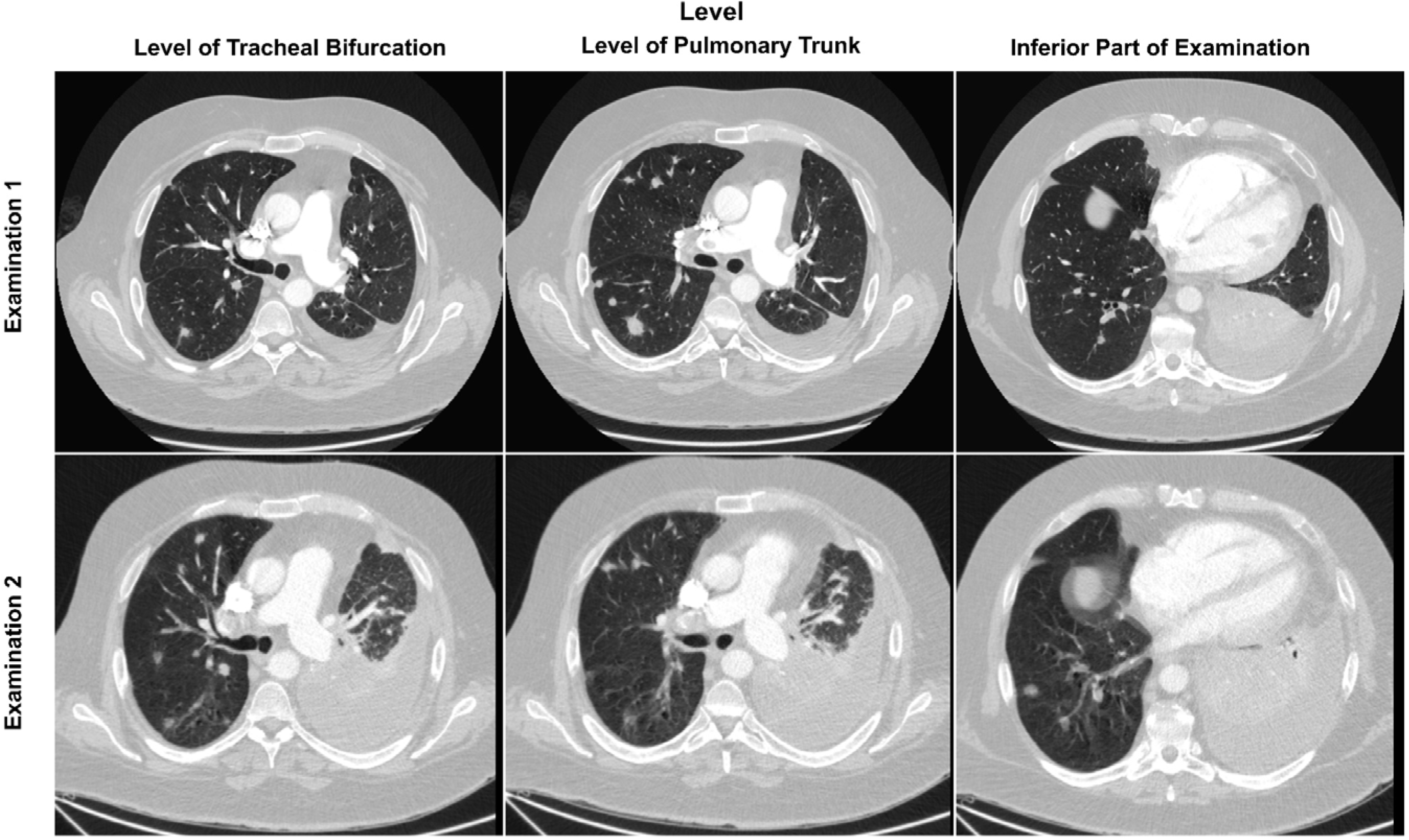
Representative examples from two CT Pulmonary Angiography (CTPA) examinations of the same patient, both having pulmonary embolism. Two CTPA examinations from the same patient within the internal dataset are presented, featuring identical window settings (width = 1500 HU, level = -400 HU) and depicted at three distinct anatomical levels. The patient exhibited pulmonary embolism in both CTPA examinations.

**Supplemental Figure 5.**
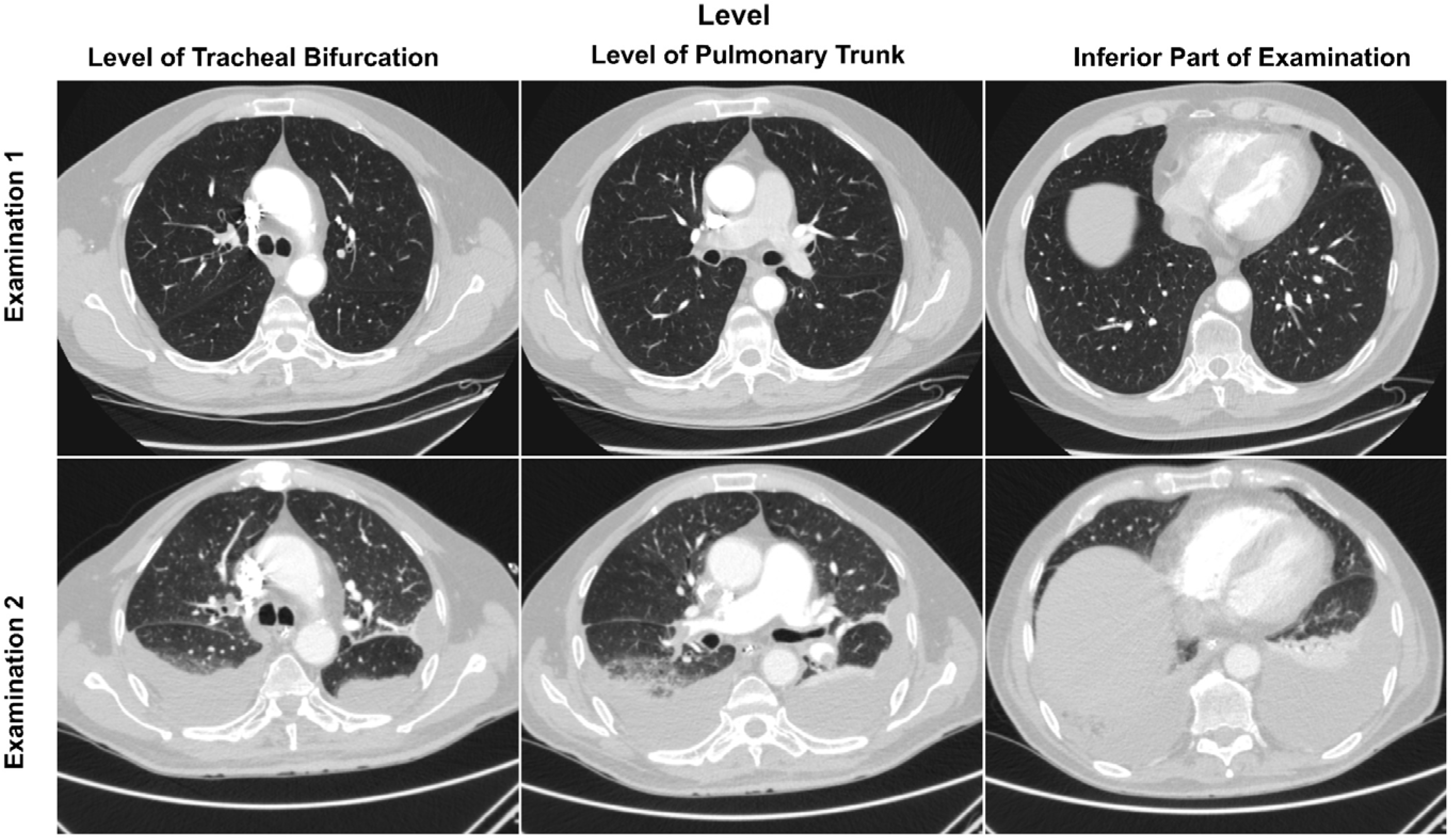
Representative examples of two CT Pulmonary Angiography (CTPA) examinations of the same patient with a pulmonary embolism in one examination but not in the other. Two CTPA examinations from the same patient within the internal dataset are presented, featuring identical window settings (width = 1500 HU, level = -400 HU) and depicted at three distinct anatomical levels. The patient exhibited pulmonary embolism in examination 2.

**Supplementary Table 1.** Diagnostic performance of the trained model without post-processing in the internal dataset.

|  | Internal Dataset (CTPAs = 679) |  |  |  |  |  |  |  |  |  |  |  |  |  |  |  |  |  |  |  |  |
| --- | --- | --- | --- | --- | --- | --- | --- | --- | --- | --- | --- | --- | --- | --- | --- | --- | --- | --- | --- | --- | --- |
|  | Test Time Augmentation Enabled |  |  |  |  |  |  |  |  |  |  |  |  |  |  |  |  |  |  |  |  |
| Metric \ Threshold | 0 | 10 | 20 | 30 | 40 | 50 | 60 | 70 | 80 | 90 | 100 | 110 | 120 | 130 | 140 | 150 | 160 | 170 | 180 | 190 | 200 |
| No. of TN | 213 | 389 | 433 | 459 | 465 | 476 | 486 | 488 | 490 | 492 | 493 | 496 | 497 | 499 | 501 | 501 | 502 | 502 | 502 | 503 | 503 |
| No. of FP | 338 | 162 | 118 | 92 | 86 | 75 | 65 | 63 | 61 | 59 | 58 | 55 | 54 | 52 | 50 | 50 | 49 | 49 | 49 | 48 | 48 |
| No. of TP | 128 | 128 | 128 | 127 | 126 | 126 | 124 | 123 | 123 | 121 | 120 | 119 | 118 | 118 | 118 | 117 | 117 | 116 | 116 | 116 | 115 |
| No. of FN | 0 | 0 | 0 | 1 | 2 | 2 | 4 | 5 | 5 | 7 | 8 | 9 | 10 | 10 | 10 | 11 | 11 | 12 | 12 | 12 | 13 |
| MCC (%) | 32.6 | 55.8 | 63.9 | 69.0 | 69.9 | 72.7 | 74.2 | 74.2 | 74.8 | 74.3 | 74.0 | 74.3 | 74.1 | 74.7 | 75.3 | 74.8 | 75.1 | 74.5 | 74.5 | 74.9 | 74.3 |
| Sensitivity (%) | 100 | 100 | 100 | 99 | 98.4 | 98.4 | 96.9 | 96.1 | 96.1 | 94.5 | 93.8 | 93.0 | 92.2 | 92.2 | 92.2 | 91.4 | 91.4 | 90.6 | 90.6 | 90.6 | 89.8 |
| Specificity (%) | 38.7 | 70.6 | 78.6 | 83.3 | 84.4 | 86.4 | 88.2 | 88.6 | 88.9 | 89.3 | 89.5 | 90.0 | 90.2 | 90.6 | 90.9 | 90.9 | 91.1 | 91.1 | 91.1 | 91.3 | 91.3 |
| Accuracy (%) | 50.2 | 76.1 | 82.6 | 86.3 | 87.0 | 88.7 | 89.8 | 90.0 | 90.3 | 90.3 | 90.3 | 90.6 | 90.6 | 90.9 | 91.2 | 91.0 | 91.2 | 91.0 | 91.0 | 91.2 | 91.0 |
| Balanced Accuracy (%) | 69.4 | 85.3 | 89.3 | 91.2 | 91.4 | 92.4 | 92.6 | 92.4 | 92.5 | 91.9 | 91.6 | 91.5 | 91.2 | 91.4 | 91.6 | 91.2 | 91.2 | 90.9 | 90.9 | 91.0 | 90.6 |
|  | Test Time Augmentation Disabled |  |  |  |  |  |  |  |  |  |  |  |  |  |  |  |  |  |  |  |  |
| No. of TN | 189 | 378 | 417 | 440 | 450 | 462 | 473 | 478 | 480 | 487 | 490 | 490 | 492 | 492 | 493 | 494 | 496 | 499 | 500 | 502 | 502 |
| No. of FP | 362 | 173 | 134 | 111 | 101 | 89 | 78 | 73 | 71 | 64 | 61 | 61 | 59 | 59 | 58 | 57 | 55 | 52 | 51 | 49 | 49 |
| No. of TP | 128 | 128 | 128 | 128 | 127 | 126 | 125 | 123 | 122 | 121 | 119 | 119 | 118 | 118 | 118 | 118 | 117 | 116 | 116 | 115 | 115 |
| No. of FN | 0 | 0 | 0 | 0 | 1 | 2 | 3 | 5 | 6 | 7 | 9 | 9 | 10 | 10 | 10 | 10 | 11 | 12 | 12 | 13 | 13 |
| MCC (%) | 29.9 | 54.0 | 60.8 | 65.4 | 67.0 | 69.2 | 71.3 | 71.5 | 71.5 | 72.8 | 72.6 | 72.6 | 72.6 | 72.6 | 72.9 | 73.2 | 73.2 | 73.6 | 73.9 | 74.0 | 74.0 |
| Sensitivity (%) | 100 | 100 | 100 | 100 | 99.2 | 98.4 | 97.7 | 96.1 | 95.3 | 94.5 | 93.0 | 93.0 | 92.2 | 92.2 | 92.2 | 92.2 | 91.4 | 90.6 | 90.6 | 89.8 | 89.8 |
| Specificity (%) | 34.3 | 68.6 | 75.7 | 79.9 | 81.7 | 83.8 | 85.8 | 86.8 | 87.1 | 88.4 | 88.9 | 88.9 | 89.3 | 89.3 | 89.5 | 89.7 | 90.0 | 90.6 | 90.7 | 91.1 | 91.1 |
| Accuracy (%) | 46.7 | 74.5 | 80.3 | 83.7 | 85.0 | 86.6 | 88.1 | 88.5 | 88.7 | 89.5 | 89.7 | 89.7 | 89.8 | 89.8 | 90.0 | 90.1 | 90.3 | 90.6 | 90.7 | 90.9 | 90.9 |
| Balanced Accuracy (%) | 67.2 | 84.3 | 87.8 | 90.0 | 90.4 | 91.1 | 91.8 | 91.4 | 91.2 | 91.4 | 91.0 | 91.0 | 90.8 | 90.8 | 90.8 | 91.0 | 90.7 | 90.6 | 90.6 | 90.4 | 90.4 |

Note. — The thresholds are in mm<sup>3</sup>. CTPAs = computed tomography (CT) pulmonary angiography (CTPA) examinations, TN = true-negative CTPAs, FP = false-positive CTPAs, TP = true-positive CTPAs, FN = false-negative CTPAs, MCC = Matthew's correlation coefficient.

**Supplementary Table 2.** Diagnostic performance of the trained model without post-processing in the external FUMPE dataset.

|  | FUMPE External Dataset (CTPAs = 34) |  |  |  |  |  |  |  |  |  |  |  |  |  |  |  |  |  |  |  |  |
| --- | --- | --- | --- | --- | --- | --- | --- | --- | --- | --- | --- | --- | --- | --- | --- | --- | --- | --- | --- | --- | --- |
|  | Test Time Augmentation Enabled |  |  |  |  |  |  |  |  |  |  |  |  |  |  |  |  |  |  |  |  |
| Metric \ Threshold | 0 | 10 | 20 | 30 | 40 | 50 | 60 | 70 | 80 | 90 | 100 | 110 | 120 | 130 | 140 | 150 | 160 | 170 | 180 | 190 | 200 |
| No. of TN | 2 | 2 | 2 | 2 | 2 | 2 | 2 | 2 | 2 | 2 | 2 | 2 | 2 | 2 | 2 | 2 | 2 | 2 | 2 | 2 | 2 |
| No. of FP | 0 | 0 | 0 | 0 | 0 | 0 | 0 | 0 | 0 | 0 | 0 | 0 | 0 | 0 | 0 | 0 | 0 | 0 | 0 | 0 | 0 |
| No. of TP | 32 | 32 | 32 | 32 | 32 | 32 | 32 | 32 | 32 | 32 | 31 | 31 | 30 | 30 | 29 | 29 | 29 | 29 | 29 | 29 | 29 |
| No. of FN | 0 | 0 | 0 | 0 | 0 | 0 | 0 | 0 | 0 | 0 | 1 | 1 | 2 | 2 | 3 | 3 | 3 | 3 | 3 | 3 | 3 |
| MCC (%) | 100.0 | 100 | 100 | 100 | 100 | 100 | 100 | 100 | 100 | 100 | 80 | 80 | 69 | 69 | 60 | 60.2 | 60.2 | 60.2 | 60.2 | 60.2 | 60.2 |
| Sensitivity (%) | 100 | 100 | 100 | 100 | 100 | 100 | 100 | 100 | 100 | 100 | 97 | 97 | 94 | 94 | 91 | 90.6 | 90.6 | 90.6 | 90.6 | 90.6 | 90.6 |
| Specificity (%) | 100 | 100 | 100 | 100 | 100 | 100 | 100 | 100 | 100 | 100 | 100 | 100 | 100 | 100 | 100 | 100 | 100 | 100 | 100 | 100 | 100 |
| Accuracy (%) | 100 | 100 | 100 | 100 | 100 | 100 | 100 | 100 | 100 | 100 | 97 | 97 | 94 | 94 | 91 | 91.2 | 91.2 | 91.2 | 91.2 | 91.2 | 91.2 |
| Balanced Accuracy (%) | 100 | 100 | 100 | 100 | 100 | 100 | 100 | 100 | 100 | 100 | 98 | 98 | 97 | 97 | 95 | 95.3 | 95.3 | 95.3 | 95.3 | 95.3 | 95.3 |
|  | Test Time Augmentation Disabled |  |  |  |  |  |  |  |  |  |  |  |  |  |  |  |  |  |  |  |  |
| No. of TN | 1 | 2 | 2 | 2 | 2 | 2 | 2 | 2 | 2 | 2 | 2 | 2 | 2 | 2 | 2 | 2 | 2 | 2 | 2 | 2 | 2 |
| No. of FP | 1 | 0 | 0 | 0 | 0 | 0 | 0 | 0 | 0 | 0 | 0 | 0 | 0 | 0 | 0 | 0 | 0 | 0 | 0 | 0 | 0 |
| No. of TP | 32 | 32 | 32 | 32 | 32 | 32 | 32 | 32 | 32 | 32 | 32 | 32 | 32 | 32 | 32 | 30 | 30 | 30 | 30 | 30 | 30 |
| No. of FN | 0 | 0 | 0 | 0 | 0 | 0 | 0 | 0 | 0 | 0 | 0 | 0 | 0 | 0 | 0 | 2 | 2 | 2 | 2 | 2 | 2 |
| MCC (%) | 69.6 | 100 | 100 | 100 | 100 | 100 | 100 | 100 | 100 | 100 | 100 | 100 | 100 | 100 | 100 | 68.5 | 68.5 | 68.5 | 68.5 | 68.5 | 68.5 |
| Sensitivity (%) | 100 | 100 | 100 | 100 | 100 | 100 | 100 | 100 | 100 | 100 | 100 | 100 | 100 | 100 | 100 | 93.8 | 93.8 | 93.8 | 93.8 | 93.8 | 93.8 |
| Specificity (%) | 50.0 | 100 | 100 | 100 | 100 | 100 | 100 | 100 | 100 | 100 | 100 | 100 | 100 | 100 | 100 | 100 | 100 | 100 | 100 | 100 | 100 |
| Accuracy (%) | 97.1 | 100 | 100 | 100 | 100 | 100 | 100 | 100 | 100 | 100 | 100 | 100 | 100 | 100 | 100 | 94.1 | 94.1 | 94.1 | 94.1 | 94.1 | 94.1 |
| Balanced Accuracy (%) | 75.0 | 100 | 100 | 100 | 100 | 100 | 100 | 100 | 100 | 100 | 100 | 100 | 100 | 100 | 100 | 96.9 | 96.9 | 96.9 | 96.9 | 96.9 | 96.9 |

Note. — The thresholds are in mm<sup>3</sup>. CTPAs = computed tomography (CT) pulmonary angiography (CTPA) examinations, TN = true-negative CTPAs, FP = false-positive CTPAs, TP = true-positive CTPAs, FN = false-negative CTPAs, MCC = Matthew's correlation coefficient. FUMPE = Ferdowsi University of Mashhad's PE dataset.

**Supplementary Table 3.**
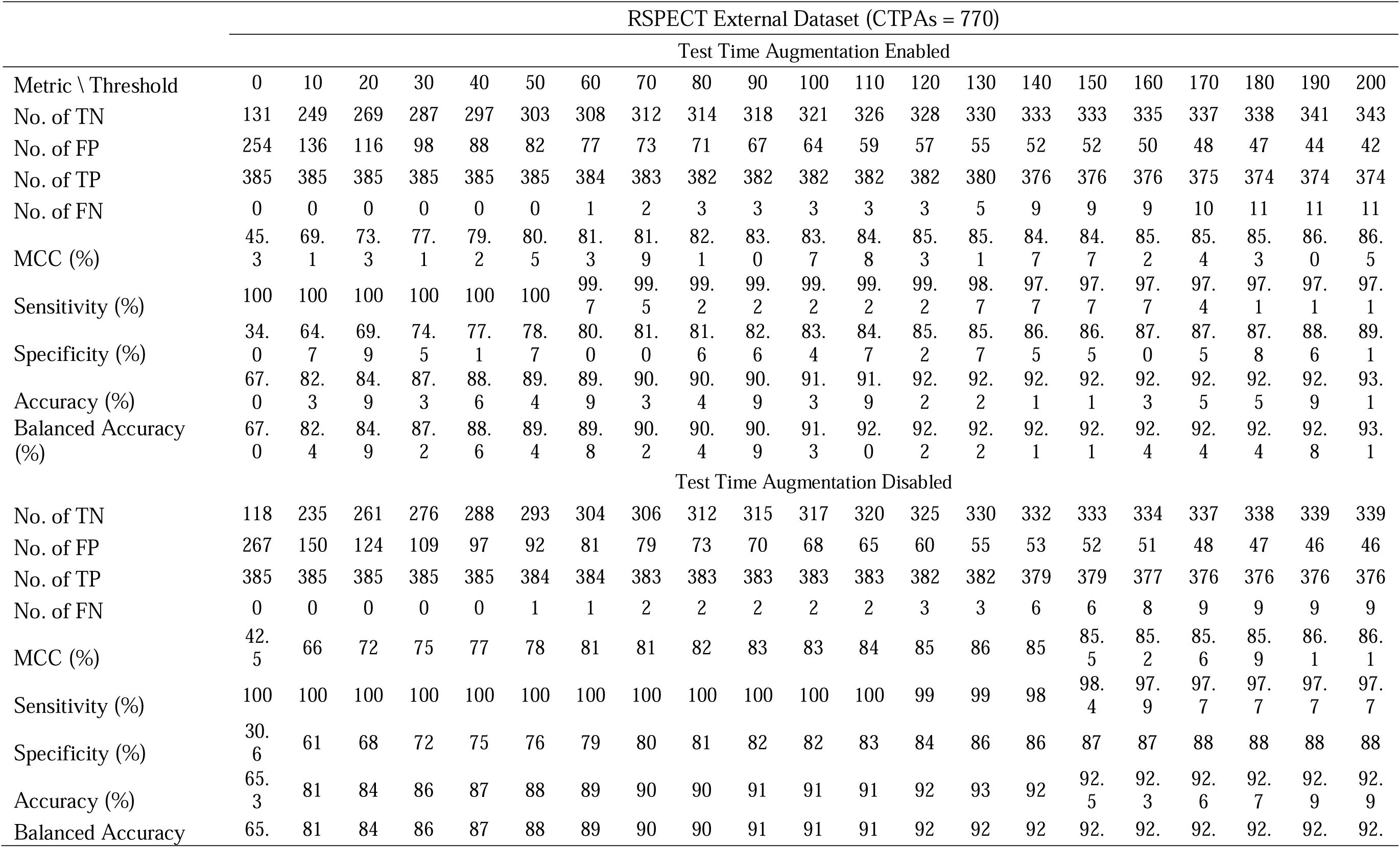
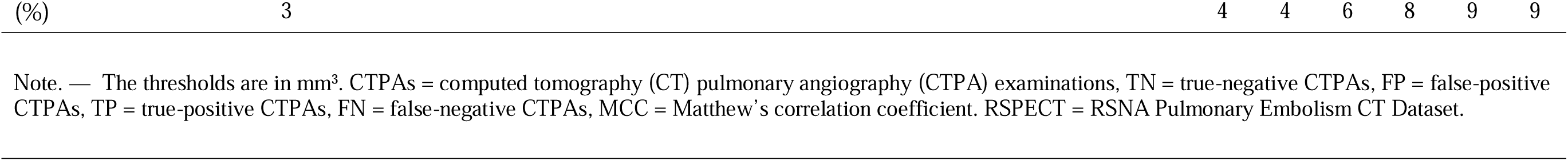
Diagnostic performance of the trained model without post-processing in the external RSPECT Dataset.

**Supplementary Table 4.**
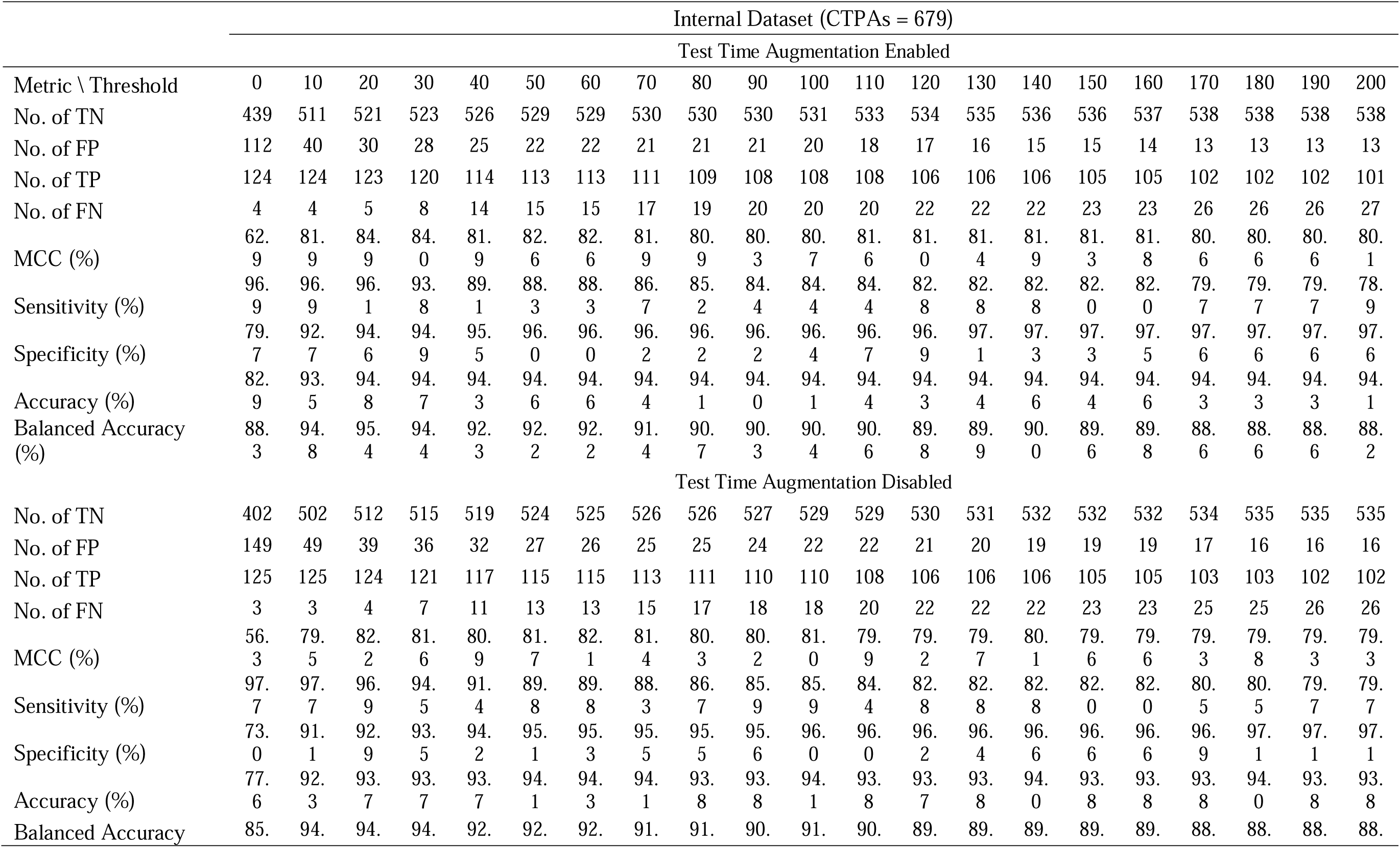
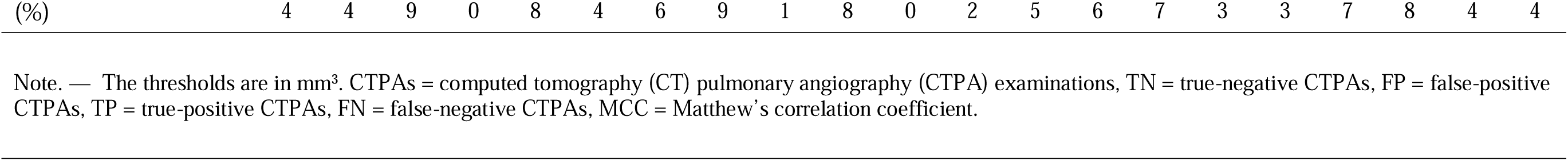
Diagnostic performance of the trained model with post-processing strategy 1 in the internal dataset.

**Supplementary Table 5.**
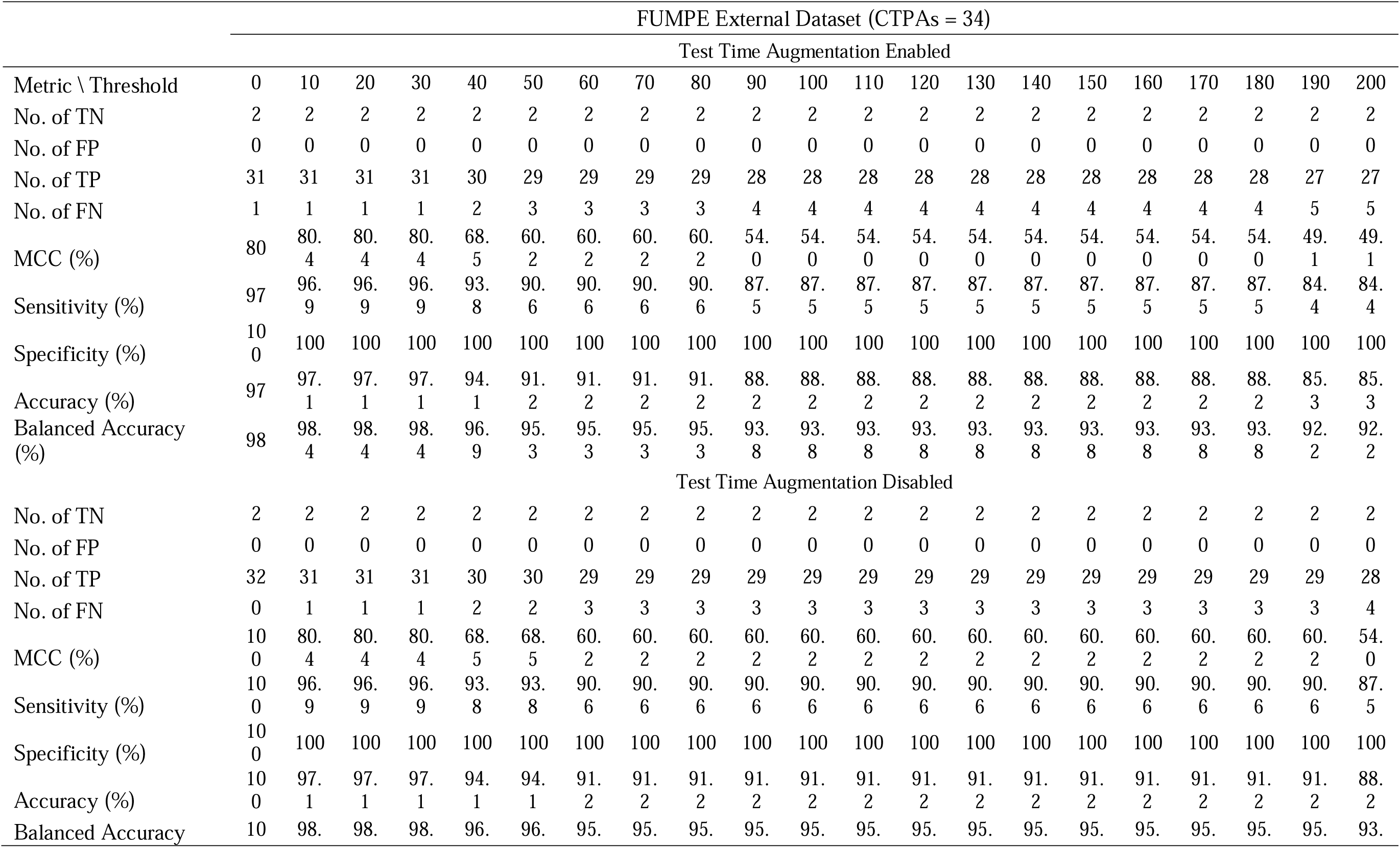
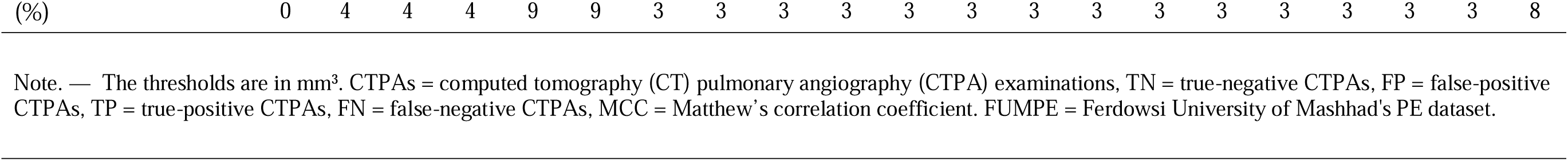
Diagnostic performance of the trained model with post-processing strategy 1 in the external FUMPE Dataset.

**Supplementary Table 6.**
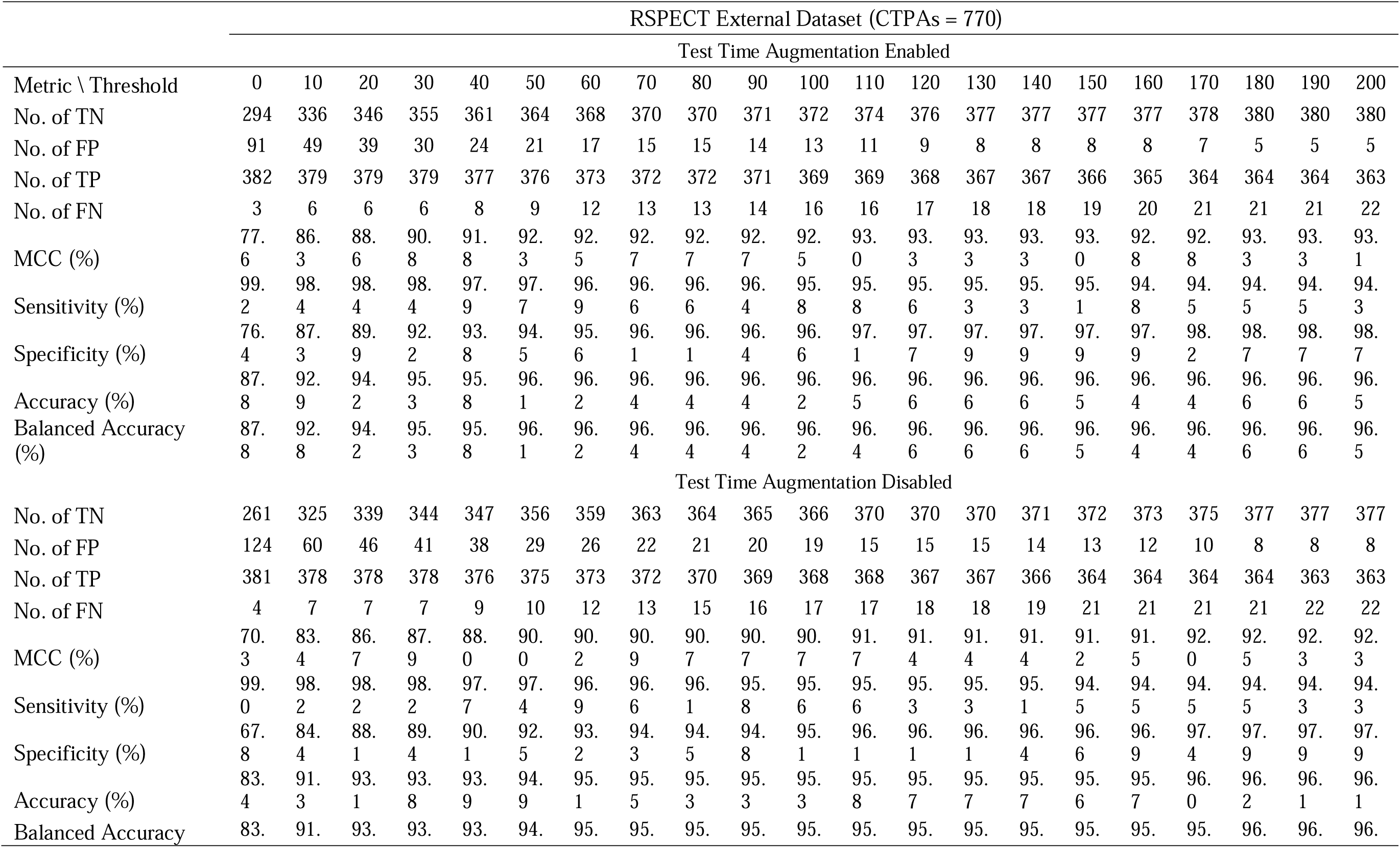
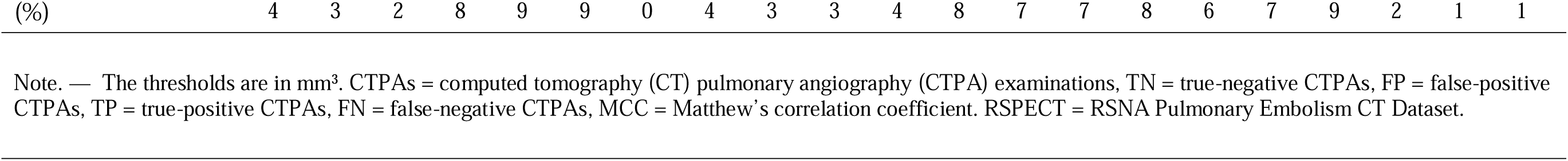
Diagnostic performance of the trained model with post-processing strategy 1 in the external RSPECT Dataset.

**Supplementary Table 7.**
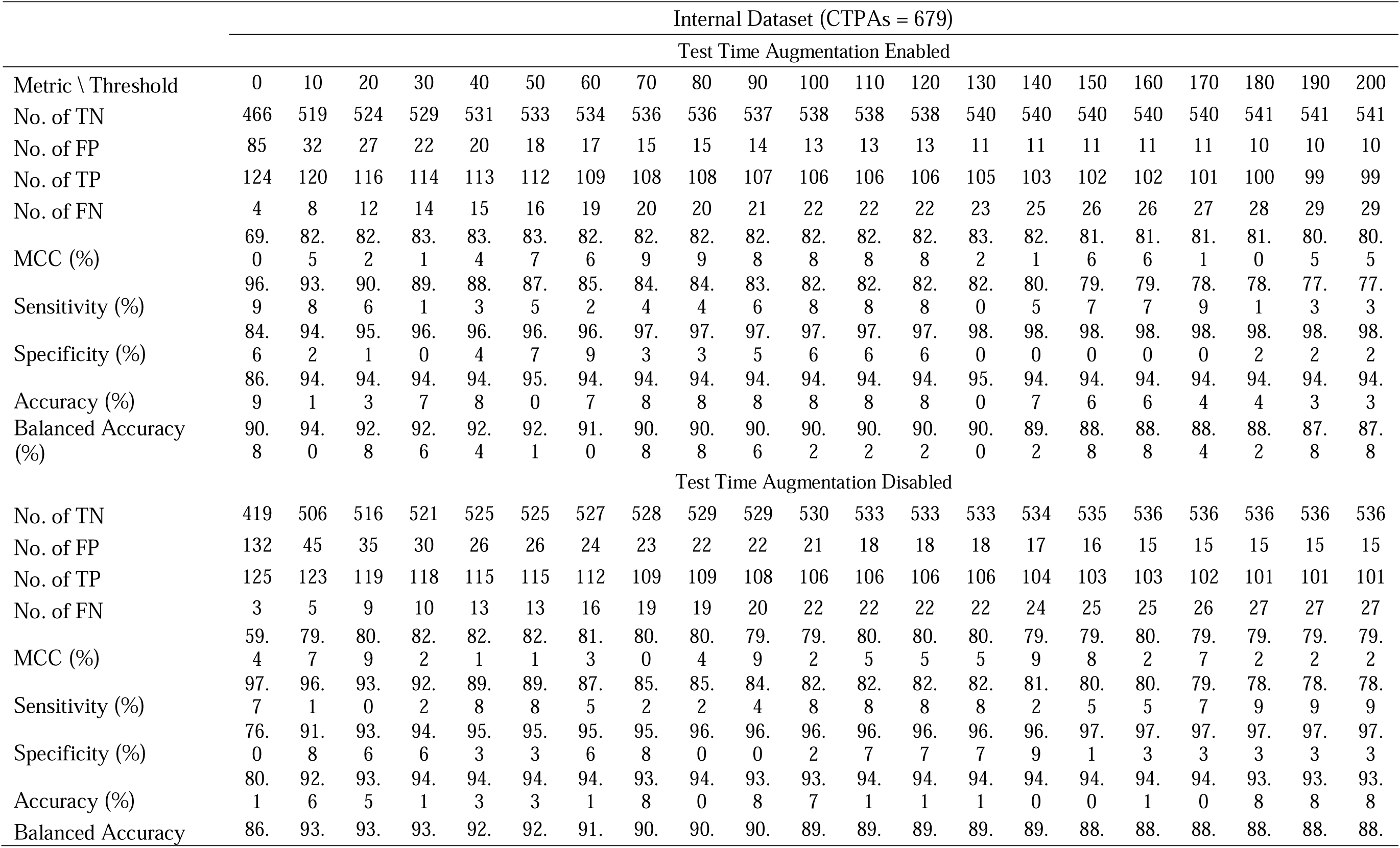
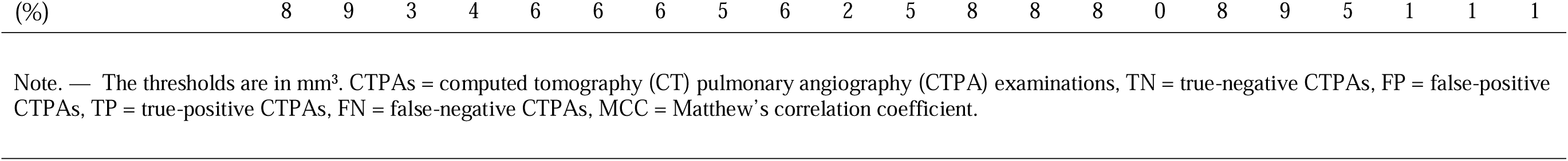
Diagnostic performance of the trained model with post-processing strategy 2 in the internal Dataset.

**Supplementary Table 8.**
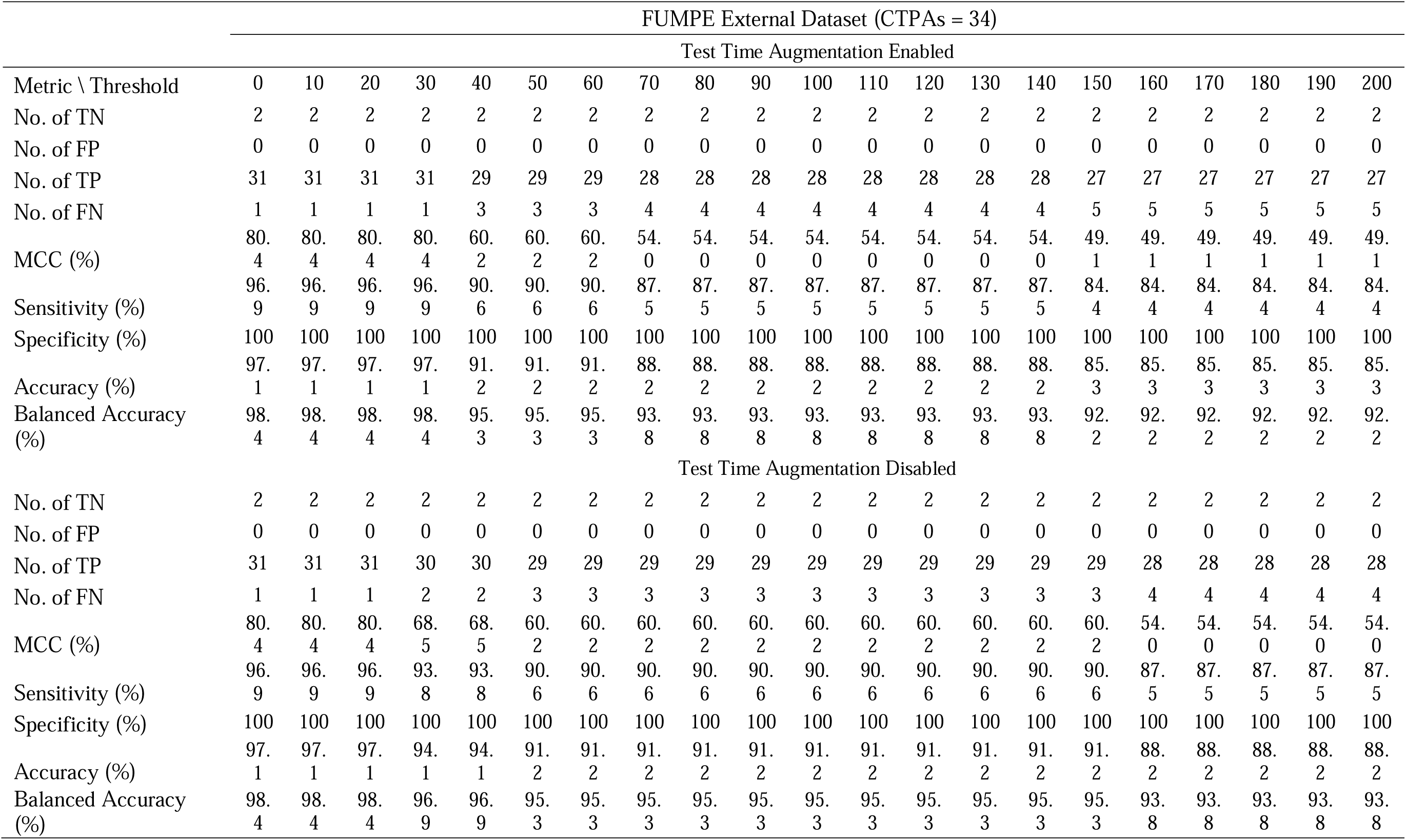
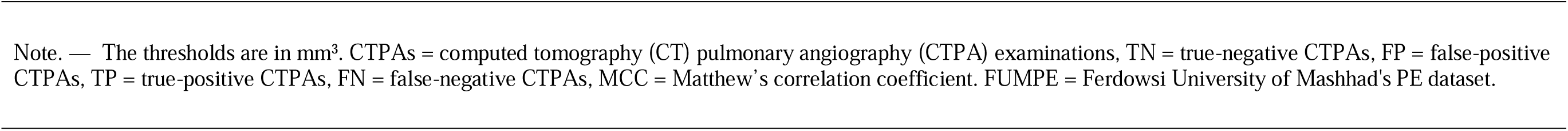
Diagnostic performance of the trained model with post-processing strategy 2 in the eternal FUMPE Dataset.

**Supplementary Table 9.**
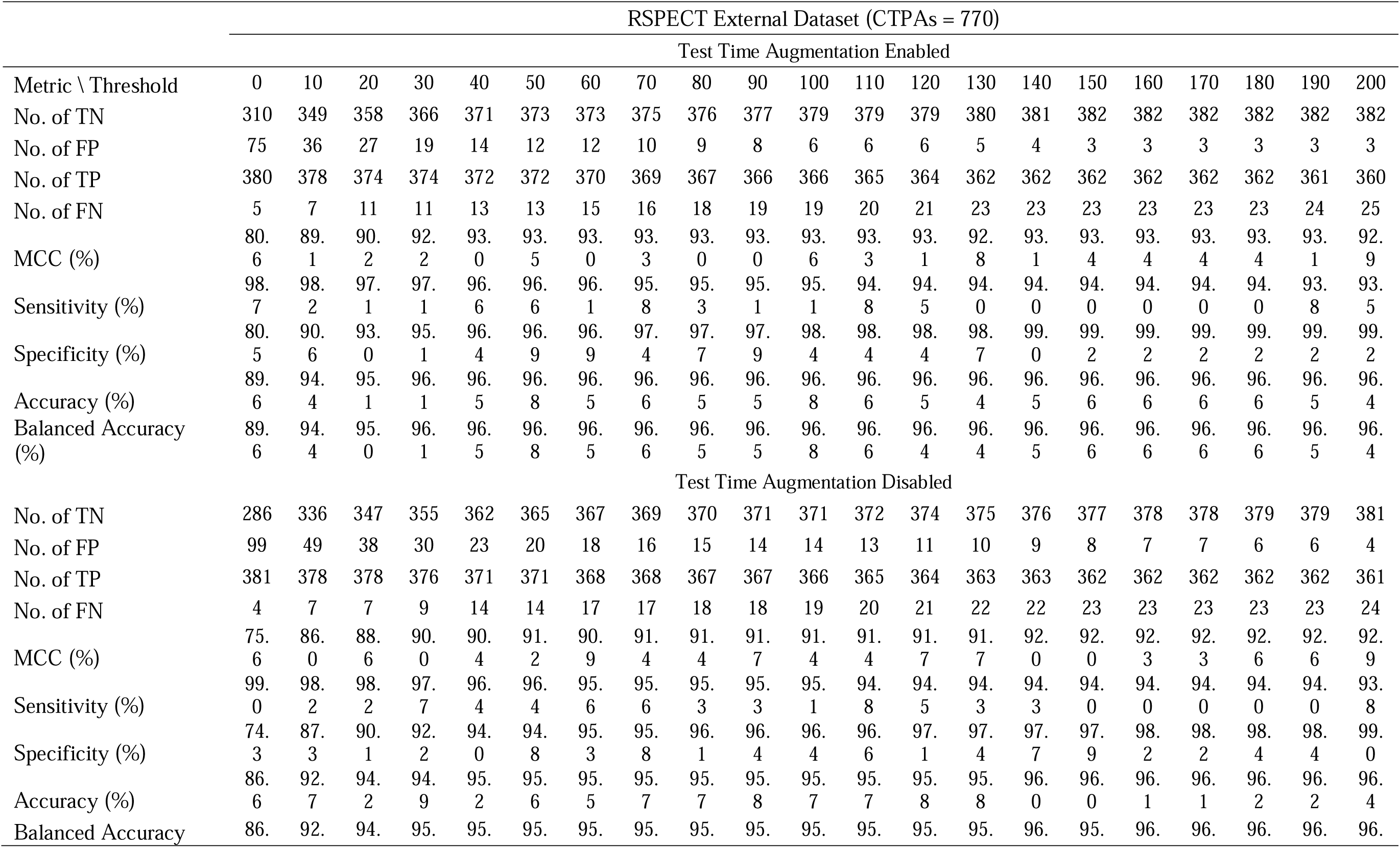
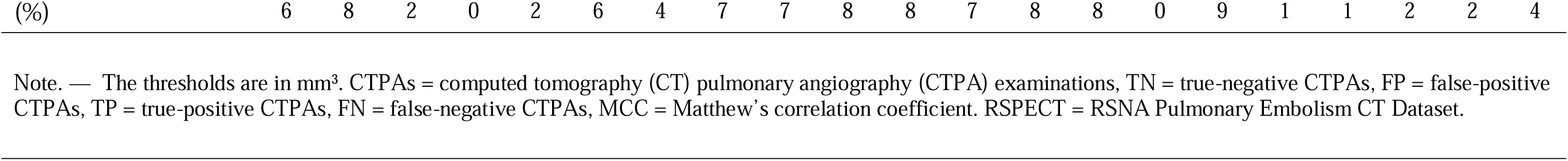
Diagnostic performance of the trained model with post-processing strategy 2 in the external RSPECT Dataset.

**Supplementary Table 10.** Diagnostic performance of the trained model without post-processing strategy in the combined testing dataset.

|  | Testing Dataset (CTPAs = 1355) |  |  |  |  |  |  |  |  |  |  |  |  |  |  |  |  |  |  |  |  |
| --- | --- | --- | --- | --- | --- | --- | --- | --- | --- | --- | --- | --- | --- | --- | --- | --- | --- | --- | --- | --- | --- |
|  | Test Time Augmentation Enabled |  |  |  |  |  |  |  |  |  |  |  |  |  |  |  |  |  |  |  |  |
| Metric \ Threshold | 0 | 10 | 20 | 30 | 40 | 50 | 60 | 70 | 80 | 90 | 100 | 110 | 120 | 130 | 140 | 150 | 160 | 170 | 180 | 190 | 200 |
| No. of TN | 346 | 640 | 704 | 748 | 764 | 781 | 796 | 802 | 806 | 812 | 816 | 824 | 827 | 831 | 836 | 836 | 839 | 841 | 842 | 846 | 848 |
| No. of FP | 592 | 298 | 234 | 190 | 174 | 157 | 142 | 136 | 132 | 126 | 122 | 114 | 111 | 107 | 102 | 102 | 99 | 97 | 96 | 92 | 90 |
| No. of TP | 417 | 417 | 417 | 417 | 417 | 417 | 416 | 415 | 414 | 414 | 413 | 413 | 412 | 410 | 405 | 405 | 405 | 404 | 403 | 403 | 403 |
| No. of FN | 0 | 0 | 0 | 0 | 0 | 0 | 1 | 2 | 3 | 3 | 4 | 4 | 5 | 7 | 12 | 12 | 12 | 13 | 14 | 14 | 14 |
| MCC (%) | 39.0 | 63.1 | 69.3 | 74.0 | 75.8 | 77.8 | 79.4 | 79.9 | 80.2 | 80.9 | 81.2 | 82.3 | 82.5 | 82.6 | 82.3 | 82.3 | 82.7 | 82.7 | 82.7 | 83.2 | 83.5 |
| Sensitivity (%) | 100 | 100 | 100 | 100 | 100 | 100 | 99.8 | 99.5 | 99.3 | 99.3 | 99.0 | 99.0 | 98.8 | 98.3 | 97.1 | 97.1 | 97.1 | 96.9 | 96.6 | 96.6 | 96.6 |
| Specificity (%) | 36.9 | 68.2 | 75.1 | 79.7 | 81.4 | 83.3 | 84.9 | 85.5 | 85.9 | 86.6 | 87.0 | 87.8 | 88.2 | 88.6 | 89.1 | 89.1 | 89.4 | 89.7 | 89.8 | 90.2 | 90.4 |
| Accuracy (%) | 56.3 | 78.0 | 82.7 | 86.0 | 87.2 | 88.4 | 89.4 | 89.8 | 90.0 | 90.5 | 90.7 | 91.3 | 91.4 | 91.6 | 91.6 | 91.6 | 91.8 | 91.9 | 91.9 | 92.2 | 92.3 |
| Balanced Accuracy (%) | 68.4 | 84.1 | 87.6 | 89.8 | 90.7 | 91.6 | 92.4 | 92.5 | 92.6 | 92.9 | 93.0 | 93.4 | 93.5 | 93.4 | 93.1 | 93.1 | 93.2 | 93.3 | 93.2 | 93.4 | 93.5 |
|  | Test Time Augmentation Disabled |  |  |  |  |  |  |  |  |  |  |  |  |  |  |  |  |  |  |  |  |
| No. of TN | 308 | 615 | 680 | 718 | 740 | 757 | 779 | 786 | 794 | 804 | 809 | 812 | 819 | 824 | 827 | 829 | 832 | 838 | 840 | 843 | 843 |
| No. of FP | 630 | 323 | 258 | 220 | 198 | 181 | 159 | 152 | 144 | 134 | 129 | 126 | 119 | 114 | 111 | 109 | 106 | 100 | 98 | 95 | 95 |
| No. of TP | 417 | 417 | 417 | 417 | 417 | 416 | 416 | 415 | 415 | 415 | 415 | 415 | 414 | 414 | 411 | 409 | 407 | 406 | 406 | 406 | 406 |
| No. of FN | 0 | 0 | 0 | 0 | 0 | 1 | 1 | 2 | 2 | 2 | 2 | 2 | 3 | 3 | 6 | 8 | 10 | 11 | 11 | 11 | 11 |
| MCC (%) | 36.2 | 60.8 | 66.9 | 70.8 | 73.1 | 74.8 | 77.3 | 78.0 | 78.9 | 80.1 | 80.8 | 81.1 | 81.8 | 82.5 | 82.3 | 82.1 | 82.1 | 82.7 | 83.0 | 83.4 | 83.4 |
| Sensitivity (%) | 100.0 | 100.0 | 100.0 | 100.0 | 100.0 | 99.8 | 99.8 | 99.5 | 99.5 | 99.5 | 99.5 | 99.5 | 99.3 | 99.3 | 98.6 | 98.1 | 97.6 | 97.4 | 97.4 | 97.4 | 97.4 |
| Specificity (%) | 32.8 | 65.6 | 72.5 | 76.5 | 78.9 | 80.7 | 83.0 | 83.8 | 84.6 | 85.7 | 86.2 | 86.6 | 87.3 | 87.8 | 88.2 | 88.4 | 88.7 | 89.3 | 89.6 | 89.9 | 89.9 |
| Accuracy (%) | 53.5 | 76.2 | 81.0 | 83.8 | 85.4 | 86.6 | 88.2 | 88.6 | 89.2 | 90.0 | 90.3 | 90.6 | 91.0 | 91.4 | 91.4 | 91.4 | 91.4 | 91.8 | 92.0 | 92.2 | 92.2 |
| Balanced Accuracy (%) | 66.4 | 82.8 | 86.2 | 88.2 | 89.4 | 90.2 | 91.4 | 91.6 | 92.0 | 92.6 | 92.8 | 93.0 | 93.3 | 93.6 | 93.4 | 93.2 | 93.2 | 93.4 | 93.5 | 93.6 | 93.6 |

Note. — The thresholds are in mm<sup>3</sup>. CTPAs = computed tomography (CT) pulmonary angiography (CTPA) examinations, TN = true-negative CTPAs, FP = false-positive CTPAs, TP = true-positive CTPAs, FN = false-negative CTPAs, MCC = Matthew's correlation coefficient. RSPECT = RSNA Pulmonary Embolism CT Dataset, FUMPE = Ferdowsi University of Mashhad's PE dataset, Testing Dataset = 551 PE negative CTPAs from internal testing set + 32 PE positive and 2 PE negative from FUMPE + 385 PE positive and 385 PE negative CTPAs from RSPECT.

**Supplementary Table 11.** Diagnostic performance of the trained model with post-processing strategy 1 in the combined testing dataset.

|  | Testing Dataset (CTPAs = 1355) |  |  |  |  |  |  |  |  |  |  |  |  |  |  |  |  |  |  |  |  |
| --- | --- | --- | --- | --- | --- | --- | --- | --- | --- | --- | --- | --- | --- | --- | --- | --- | --- | --- | --- | --- | --- |
|  | Test Time Augmentation Enabled |  |  |  |  |  |  |  |  |  |  |  |  |  |  |  |  |  |  |  |  |
| Metric \ Threshold | 0 | 10 | 20 | 30 | 40 | 50 | 60 | 70 | 80 | 90 | 100 | 110 | 120 | 130 | 140 | 150 | 160 | 170 | 180 | 190 | 200 |
| No. of TN | 735 | 849 | 869 | 880 | 889 | 895 | 899 | 902 | 902 | 903 | 905 | 909 | 912 | 914 | 915 | 915 | 916 | 918 | 920 | 920 | 920 |
| No. of FP | 203 | 89 | 69 | 58 | 49 | 43 | 39 | 36 | 36 | 35 | 33 | 29 | 26 | 24 | 23 | 23 | 22 | 20 | 18 | 18 | 18 |
| No. of TP | 413 | 410 | 410 | 410 | 407 | 405 | 402 | 401 | 401 | 399 | 397 | 397 | 396 | 395 | 395 | 394 | 393 | 392 | 392 | 391 | 390 |
| No. of FN | 4 | 7 | 7 | 7 | 10 | 12 | 15 | 16 | 16 | 18 | 20 | 20 | 21 | 22 | 22 | 23 | 24 | 25 | 25 | 26 | 27 |
| MCC (%) | 71.7 | 85.0 | 87.8 | 89.4 | 90.2 | 90.8 | 90.9 | 91.2 | 91.2 | 91.0 | 90.9 | 91.6 | 91.9 | 92.0 | 92.2 | 92.0 | 92.0 | 92.2 | 92.5 | 92.3 | 92.2 |
| Sensitivity (%) | 99.0 | 98.3 | 98.3 | 98.3 | 97.6 | 97.1 | 96.4 | 96.2 | 96.2 | 95.7 | 95.2 | 95.2 | 95.0 | 94.7 | 94.7 | 94.5 | 94.2 | 94.0 | 94.0 | 93.8 | 93.5 |
| Specificity (%) | 78.4 | 90.5 | 92.6 | 93.8 | 94.8 | 95.4 | 95.8 | 96.2 | 96.2 | 96.3 | 96.5 | 96.9 | 97.2 | 97.4 | 97.5 | 97.5 | 97.7 | 97.9 | 98.1 | 98.1 | 98.1 |
| Accuracy (%) | 84.7 | 92.9 | 94.4 | 95.2 | 95.6 | 95.9 | 96.0 | 96.2 | 96.2 | 96.1 | 96.1 | 96.4 | 96.5 | 96.6 | 96.7 | 96.6 | 96.6 | 96.7 | 96.8 | 96.8 | 96.7 |
| Balanced Accuracy (%) | 88.7 | 94.4 | 95.4 | 96.0 | 96.2 | 96.2 | 96.1 | 96.2 | 96.2 | 96.0 | 95.8 | 96.0 | 96.1 | 96.0 | 96.1 | 96.0 | 95.9 | 95.9 | 96.0 | 95.9 | 95.8 |
|  | Test Time Augmentation Disabled |  |  |  |  |  |  |  |  |  |  |  |  |  |  |  |  |  |  |  |  |
| No. of TN | 665 | 829 | 853 | 861 | 868 | 882 | 886 | 891 | 892 | 894 | 897 | 901 | 902 | 903 | 905 | 906 | 907 | 911 | 914 | 914 | 914 |
| No. of FP | 273 | 109 | 85 | 77 | 70 | 56 | 52 | 47 | 46 | 44 | 41 | 37 | 36 | 35 | 33 | 32 | 31 | 27 | 24 | 24 | 24 |
| No. of TP | 413 | 409 | 409 | 409 | 406 | 405 | 402 | 401 | 399 | 398 | 397 | 397 | 396 | 396 | 395 | 393 | 393 | 393 | 393 | 392 | 391 |
| No. of FN | 4 | 8 | 8 | 8 | 11 | 12 | 15 | 16 | 18 | 19 | 20 | 20 | 21 | 21 | 22 | 24 | 24 | 24 | 24 | 25 | 26 |
| MCC (%) | 64.6 | 82.1 | 85.4 | 86.5 | 86.9 | 88.8 | 88.8 | 89.4 | 89.2 | 89.3 | 89.6 | 90.3 | 90.3 | 90.4 | 90.6 | 90.4 | 90.5 | 91.2 | 91.7 | 91.5 | 91.3 |
| Sensitivity (%) | 99.0 | 98.1 | 98.1 | 98.1 | 97.4 | 97.1 | 96.4 | 96.2 | 95.7 | 95.4 | 95.2 | 95.2 | 95.0 | 95.0 | 94.7 | 94.2 | 94.2 | 94.2 | 94.2 | 94.0 | 93.8 |
| Specificity (%) | 70.9 | 88.4 | 90.9 | 91.8 | 92.5 | 94.0 | 94.5 | 95.0 | 95.1 | 95.3 | 95.6 | 96.1 | 96.2 | 96.3 | 96.5 | 96.6 | 96.7 | 97.1 | 97.4 | 97.4 | 97.4 |
| Accuracy (%) | 79.6 | 91.4 | 93.1 | 93.7 | 94.0 | 95.0 | 95.1 | 95.4 | 95.3 | 95.4 | 95.5 | 95.8 | 95.8 | 95.9 | 95.9 | 95.9 | 95.9 | 96.2 | 96.5 | 96.4 | 96.3 |
| Balanced Accuracy (%) | 84.9 | 93.2 | 94.5 | 94.9 | 94.9 | 95.6 | 95.4 | 95.6 | 95.4 | 95.4 | 95.4 | 95.6 | 95.6 | 95.6 | 95.6 | 95.4 | 95.4 | 95.6 | 95.8 | 95.7 | 95.6 |

Note. — The thresholds are in mm<sup>3</sup>. CTPAs = computed tomography (CT) pulmonary angiography (CTPA) examinations, TN = true-negative CTPAs, FP = false-positive CTPAs, TP = true-positive CTPAs, FN = false-negative CTPAs, MCC = Matthew's correlation coefficient. RSPECT = RSNA Pulmonary Embolism CT Dataset, FUMPE = Ferdowsi University of Mashhad's PE dataset, Testing Dataset = 551 PE negative CTPAs from internal testing set + 32 PE positive and 2 PE negative from FUMPE + 385 PE positive and 385 PE negative CTPAs from RSPECT.

**Supplementary Table 12.** Diagnostic performance of the trained model with post-processing strategy 2 in the combined testing dataset.

|  | Testing Dataset (CTPAs = 1355) |  |  |  |  |  |  |  |  |  |  |  |  |  |  |  |  |  |  |  |  |
| --- | --- | --- | --- | --- | --- | --- | --- | --- | --- | --- | --- | --- | --- | --- | --- | --- | --- | --- | --- | --- | --- |
|  | Test Time Augmentation Enabled |  |  |  |  |  |  |  |  |  |  |  |  |  |  |  |  |  |  |  |  |
| Metric \ Threshold | 0 | 10 | 20 | 30 | 40 | 50 | 60 | 70 | 80 | 90 | 100 | 110 | 120 | 130 | 140 | 150 | 160 | 170 | 180 | 190 | 200 |
| No. of TN | 778 | 870 | 884 | 897 | 904 | 908 | 909 | 913 | 914 | 916 | 919 | 919 | 919 | 922 | 923 | 924 | 924 | 924 | 925 | 925 | 925 |
| No. of FP | 160 | 68 | 54 | 41 | 34 | 30 | 29 | 25 | 24 | 22 | 19 | 19 | 19 | 16 | 15 | 14 | 14 | 14 | 13 | 13 | 13 |
| No. of TP | 411 | 409 | 405 | 405 | 401 | 401 | 399 | 397 | 395 | 394 | 394 | 393 | 392 | 390 | 390 | 389 | 389 | 389 | 389 | 388 | 387 |
| No. of FN | 6 | 8 | 12 | 12 | 16 | 16 | 18 | 20 | 22 | 23 | 23 | 24 | 25 | 27 | 27 | 28 | 28 | 28 | 28 | 29 | 30 |
| MCC (%) | 76.2 | 87.8 | 89.1 | 91.1 | 91.5 | 92.1 | 91.9 | 92.2 | 92.0 | 92.2 | 92.7 | 92.5 | 92.4 | 92.5 | 92.7 | 92.7 | 92.7 | 92.7 | 92.9 | 92.7 | 92.5 |
| Sensitivity (%) | 98.6 | 98.1 | 97.1 | 97.1 | 96.2 | 96.2 | 95.7 | 95.2 | 94.7 | 94.5 | 94.5 | 94.2 | 94.0 | 93.5 | 93.5 | 93.3 | 93.3 | 93.3 | 93.3 | 93.0 | 92.8 |
| Specificity (%) | 82.9 | 92.8 | 94.2 | 95.6 | 96.4 | 96.8 | 96.9 | 97.3 | 97.4 | 97.7 | 98.0 | 98.0 | 98.0 | 98.3 | 98.4 | 98.5 | 98.5 | 98.5 | 98.6 | 98.6 | 98.6 |
| Accuracy (%) | 87.7 | 94.4 | 95.1 | 96.1 | 96.3 | 96.6 | 96.5 | 96.7 | 96.6 | 96.7 | 96.9 | 96.8 | 96.8 | 96.8 | 96.9 | 96.9 | 96.9 | 96.9 | 97.0 | 96.9 | 96.8 |
| Balanced Accuracy (%) | 90.8 | 95.4 | 95.6 | 96.4 | 96.3 | 96.5 | 96.3 | 96.2 | 96.0 | 96.1 | 96.2 | 96.1 | 96.0 | 95.9 | 96.0 | 95.9 | 95.9 | 95.9 | 96.0 | 95.8 | 95.7 |
|  | Test Time Augmentation Disabled |  |  |  |  |  |  |  |  |  |  |  |  |  |  |  |  |  |  |  |  |
| No. of TN | 707 | 844 | 865 | 878 | 889 | 892 | 896 | 899 | 901 | 902 | 903 | 907 | 909 | 910 | 912 | 914 | 916 | 916 | 917 | 917 | 919 |
| No. of FP | 231 | 94 | 73 | 60 | 49 | 46 | 42 | 39 | 37 | 36 | 35 | 31 | 29 | 28 | 26 | 24 | 22 | 22 | 21 | 21 | 19 |
| No. of TP | 412 | 409 | 409 | 406 | 401 | 400 | 397 | 397 | 396 | 396 | 395 | 394 | 393 | 392 | 392 | 391 | 390 | 390 | 390 | 390 | 389 |
| No. of FN | 5 | 8 | 8 | 11 | 16 | 17 | 20 | 20 | 21 | 21 | 22 | 23 | 24 | 25 | 25 | 26 | 27 | 27 | 27 | 27 | 28 |
| MCC (%) | 68.6 | 84.1 | 87.1 | 88.4 | 89.1 | 89.4 | 89.5 | 90.0 | 90.1 | 90.3 | 90.2 | 90.7 | 90.9 | 90.8 | 91.2 | 91.3 | 91.5 | 91.5 | 91.7 | 91.7 | 91.8 |
| Sensitivity (%) | 98.8 | 98.1 | 98.1 | 97.4 | 96.2 | 95.9 | 95.2 | 95.2 | 95.0 | 95.0 | 94.7 | 94.5 | 94.2 | 94.0 | 94.0 | 93.8 | 93.5 | 93.5 | 93.5 | 93.5 | 93.3 |
| Specificity (%) | 75.4 | 90.0 | 92.2 | 93.6 | 94.8 | 95.1 | 95.5 | 95.8 | 96.1 | 96.2 | 96.3 | 96.7 | 96.9 | 97.0 | 97.2 | 97.4 | 97.7 | 97.7 | 97.8 | 97.8 | 98.0 |
| Accuracy (%) | 82.6 | 92.5 | 94.0 | 94.8 | 95.2 | 95.4 | 95.4 | 95.6 | 95.7 | 95.8 | 95.8 | 96.0 | 96.1 | 96.1 | 96.2 | 96.3 | 96.4 | 96.4 | 96.5 | 96.5 | 96.5 |
| Balanced Accuracy (%) | 87.1 | 94.0 | 95.2 | 95.5 | 95.5 | 95.5 | 95.4 | 95.5 | 95.6 | 95.6 | 95.5 | 95.6 | 95.5 | 95.5 | 95.6 | 95.6 | 95.6 | 95.6 | 95.6 | 95.6 | 95.6 |

Note. — The thresholds are in mm<sup>3</sup>. CTPAs = computed tomography (CT) pulmonary angiography (CTPA) examinations, TN = true-negative CTPAs, FP = false-positive CTPAs, TP = true-positive CTPAs, FN = false-negative CTPAs, MCC = Matthew's correlation coefficient. RSPECT = RSNA Pulmonary Embolism CT Dataset, FUMPE = Ferdowsi University of Mashhad's PE dataset, Testing Dataset = 551 PE negative CTPAs from internal testing set + 32 PE positive and 2 PE negative from FUMPE + 385 PE positive and 385 PE negative CTPAs from RSPECT.

